# Adult dengue vaccination in Singapore: A modelling study to inform policy

**DOI:** 10.1101/2025.10.01.25337064

**Authors:** Abhishek Senapati, Tun-Linn Thein, Yee-Sin Leo, Hannah Eleanor Clapham

## Abstract

Dengue virus, with all four serotypes in cocirculation, has created significant epidemiological and economic burdens in Singapore. Despite integrated vector control programs, the magnitude and frequency of dengue outbreaks has increased over the last few decades, which highlights the limits of existing strategies. In this context, vaccination has emerged as a promising approach to enhance population immunity and complement ongoing efforts. Qdenga (TAK-003), a commercially available dengue vaccine, has been recommended by the the World Health Organization Strategic Advisory Group of Experts (WHO-SAGE) for use in high-transmission settings. Although classified as a low transmission setting, Singapore faces a recurring public health burden from dengue, with high costs associated with both healthcare and sustained vector control. The age distribution of dengue in Singapore differs from high transmission settings, with more cases in young adults and hospitalizations concentrated among older individuals. These factors, combined with the complex efficacy profile of Qdenga, varying by baseline serostatus, infecting serotype, make it challenging to directly apply global vaccination recommendations to Singapore. Therefore, the population-level impact of introducing dengue vaccination remains uncertain, underscoring the need for context-specific evaluation.

To estimate the potential public health impact of introducing routine dengue vaccination in Singapore, we develop an age-stratified, multi-serotype, compartmental transmission model informed by age-specific dengue seroprevalence and routine surveillance data. Our model predicts that vaccination can avert up to on average 9%, 12%, 7%, and 5% cases in DENV-1–4 dominant serotype scenarios respectively, over a 10-year routine vaccination program with 80% vaccine coverage. Moreover, dengue hospitalizations can be reduced by 12%–15% across all dominant serotype scenarios. Our results suggest that, in Singapore, targeting older age groups will be more beneficial than the 6–16-year window recommended by the WHO-SAGE for high-transmission settings. Vaccinating individuals aged 17–30 years achieves the greatest reduction in cases, whereas targeting those aged 51–70 years leads to the highest reduction in hospitalizations.

Our model-based analysis provides useful insights to support policymakers and public health authorities in designing evidence-based, dengue vaccination strategies in Singapore. The findings also underscore the importance of tailoring dengue vaccination programs to local epidemiological conditions for effective disease control.

**Authors summary:** Singapore is experiencing more frequent dengue outbreaks, which impose a substantial burden through treatment costs and sustained vector control programs. Vaccination is a key public health measure that can help ease this burden by complementing existing control efforts. In clinical trials, Qdenga (TAK-003) demonstrated consistent protections against DENV-1 and DENV-2 among vaccine recipients irrespective of their baseline serostatus, but uncertainty remains for DENV-3 and DENV-4. Although WHO-SAGE recommended its use among children aged 6–16 years in high dengue transmission setting, uncertainty arises when extrapolating these recommendations to low transmission settings like Singapore. Moreover, unlike high transmission settings, Singapore currently shows a different age pattern of dengue burden, with higher incidence in young adults and more severe outcomes among older individuals, highlighting the need to contextualize these global recommendations using country-specific data and modelling approaches.

To address this, we developed a detailed modelling framework that integrates local epidemiological characteristics and operational aspects of vaccine implementation and evaluated the potential impact of introducing a routine vaccination program in Singapore. We show that, with 80% coverage, vaccination can avert up to 12% of reported cases and 15% of hospitalizations over a 10-year period, with impact varying mainly by circulating serotype and target age group. Our findings suggest that in low-transmission settings like Singapore, vaccinating adults will achieve a greater public health impact than targeting children, as currently recommended for high-transmission settings by WHO-SAGE. These findings provide timely evidence to support national policy discussions on dengue vaccination in Singapore and other low-transmission settings, where adult disease burden is substantial.

## Introduction

Dengue fever, a mosquito-borne viral infection caused by the dengue virus, poses a significant global health burden. Recent studies estimated more than 60 million symptomatic dengue infections occurs annually, with over half the global population at risk, predominantly in tropical and subtropical regions (1,2). Globally, the burden of dengue has increased exponentially over the past few decades due to urbanization, globalization, and climate change, which enhances mosquito breeding conditions. The epidemiology of dengue is complex, driven by four virus serotypes (DENV-1-4), each contributing to varying degrees of disease severity and immunity. The primary infection caused by any of the four serotypes typically causes no or mild disease and provides long-term homologous protection and temporary heterologous protection to the infected individuals (3). On the other hand, secondary dengue infection is thought to be responsible for increased risk of severe disease due to antibody-dependent enhancement (ADE) (4). The subsequent third and fourth dengue infections are rarely observed to have severe outcomes (5).

The dengue transmission intensity in Singapore is quite low compared to other Southeast Asian countries, mainly due to its successful integrated vector control and robust disease surveillance programs (6–9). Despite such control efforts, Singapore has experienced frequent and larger outbreaks in recent decades. Although the exact drivers behind these outbreaks remain uncertain, they may be attributed to factors such as low-population immunity due to successful control. In addition to the public health burden, dengue imposes significant economic burden due to exceptionally high treatment expenses and productivity losses from work absenteeism in Singapore (9,10). The ongoing transmission highlights the limitation of existing vector control programs and therefore dengue vaccine emerges as a promising tool that could complement the ongoing dengue control, efforts, including Wolbachia, by boosting population immunity in Singapore.

The development of dengue vaccines has been an active area of research, with two vaccines: CYD-TDV (Dengvaxia) and TAK-003 (Qdenga) are commercially available. Another vaccine candidate, TV-003, has completed Phase III clinical trials but has not yet been recommended for use (11), and a new trial has recently been initiated in Asia. The first licensed vaccine, Dengvaxia, developed by Sanofi Pasteur, was approved in several dengue-endemic countries, including Singapore. However, due to its safety and efficacy being limited to individuals with prior dengue exposure, pre-vaccination screening was required. These constraints, along with safety concerns, led to low vaccine uptake, and Sanofi has announced plans to discontinue its production. On the other hand, the next commercially available vaccine Qdenga, developed by Takeda, has been recommended by WHO-SAGE for use among individuals aged 6–16 years in high dengue transmission settings, following a two-dose schedule administered three months apart, without the need for pre-vaccination screening (12,13). Qdenga has been licensed in several dengue-endemic countries, including Brazil, Indonesia, and Thailand, but has not yet been licensed for use in Singapore.

Clinical trial data from dengue endemic regions of Latin America and Asia have shown that the efficacy of Qdenga varies significantly with respect to baseline serostatus of the vaccinated population and infecting dengue serotypes (14–18). The long-term efficacy data (approximately 57 months after the first dose of vaccine) showed a higher level of protection against virologically confirmed dengue (VCD) with DENV-1 and 2, among both seropositive and seronegative individuals. The vaccine provided better protection to seropositive individuals, although the protection was incomplete, and no significant protection was observed in seronegative individuals against VCD caused by DENV-3 and DENV-4. The vaccine is well protective of seropositive individuals against hospitalization due to DENV-1, 2, and 3. While it protected seronegative individuals from getting hospitalized due to DENV-1, it failed to protect them from DENV-3. It is not possible to estimate the efficacy against hospitalization due to DENV-4 in both seropositive and seronegative individuals, and due to DENV-2 in seronegative individuals, as there were insufficient hospitalization events for these groups in the clinical trials (18).

It is important to note that all clinical trials of Qdenga have been conducted in high dengue transmission settings, and current WHO-SAGE recommendations are tailored to such contexts. In contrast, Singapore has a unique dengue epidemiology characterized by relatively low transmission intensity, and the co-circulation of all four serotypes with periodic shifts in dominance. Given these distinct features and the variability and uncertainty in vaccine efficacy across serotypes and prior exposure, it remains unclear what the overall population-level impact of vaccination would be in Singapore. Furthermore, it is uncertain whether the current WHO-SAGE recommendations, particularly regarding the target age group, would be appropriate in this low-transmission setting.

In this study, we develop an age-stratified, multi-serotype compartmental transmission model to evaluate the potential impact of dengue vaccination strategies in Singapore. The model incorporates local transmission dynamics, population immunity, and serotype- and serostatus-specific efficacy data for Qdenga, based on clinical trial findings. With our modelling framework, we project the population level impact in terms of cases and hospitalization averted due to the introduction of a 10-year routine vaccination program in Singapore across different dominant serotype scenarios and assesses the effectiveness of various age-targeting strategies.

## Methods

### Transmission model

We developed a deterministic, four-serotype, age-stratified compartmental transmission model that includes both the human and adult mosquito population explicitly (see Eq. S1–S6 and Table S1). Humans are assumed to be born susceptible and can get primary infection from one of the four dengue serotypes: DENV-1-4. Upon recovery from primary infection, they acquire permanent protection against homologous infection and transient cross-protection against heterologous infection. Once the cross-protection stage is over, individuals become susceptible to secondary infection by all the serotypes other than their primary infection causing serotype. Based on the rarity of third and fourth infections (5), we make the assumption that individuals acquire complete protection from dengue infection after secondary infection (19,20). To account for ADE, we model secondary infections as being more likely to result in symptomatic cases and hospitalizations compared to primary infections. We assume that the fraction of dengue infections to be reported as cases (i.e., reporting rate) and the fraction of cases required hospitalization (i.e., hospitalization rate) depend on both age and individual’s pre-exposure history (i.e., whether it is a primary or secondary infection). We estimated the reporting rate from the age-stratified annual dengue incidence of cases per 100000 population in Singapore. The hospitalization rates were calculated from the data published in a previous study (21).

We also consider the mosquito population explicitly in the model and it is assumed to be constant over time. Mosquitoes become infected upon feeding on humans infected by a specific serotype and subsequently infect susceptible humans. Due to their short life span they remain infected for the rest of their life.

To model vaccine rollout program, we consider that the vaccination is introduced among all the individuals except those who are currently infected, within a targeted age group with a given vaccine coverage. Disease progression among vaccinated individuals will follow the similar transitions as in unvaccinated individuals, but with modified rate of symptomatic disease and hospitalization based on the available vaccine efficacy data of Qdenga. Since vaccine efficacy estimates from vaccine trials showed variability mainly with respect to infecting serotypes and baseline serostatus of vaccinated individuals (see Fig S1), we stratify efficacy by these two factors. Although the vaccine trials were conducted in individuals aged 4–16 years, we assume similar efficacy for older age groups, based on findings from an immunobridging study demonstrating comparable immunogenicity in adults (22).

### Model parametrization

We consider only Singaporean citizen and permanent resident (PR) in our modelling study as the age-stratified sero-survey and surveillance data are reported only for these groups. We divide the human population into 91 age groups: 0 to 89 years with 1 year age bracket and 90 years & above. The historical data for age distribution among population, birth and death rate in Singapore are taken from Singapore Department of Statistics (23) and the projection of demographic trends are taken from World Population Prospects (WPP)(24).

We used a sero-catalytic model on the age-specific seroprevalence data from the sero-survey conducted in 2013 in Singapore (25) to estimate the dengue pre-exposure history, i.e., proportion of individuals who had exactly one infection, at least one infection, and two or more infections in each age group (see Fig S2). We further stratified the exposure history according to serotype based on serotype distribution of historical reported dengue cases data (26) as well as serology data (27). We considered Singapore population experienced the highest exposure against DENV-2, followed by DENV-1, 3, and 4. These estimates have been translated as initial conditions of the dengue transmission model. The mean annual force of infection estimated using the catalytic model was 0.01, which aligns with the previous study (25). We set the serotype specific infected mosquito to human transmission parameters (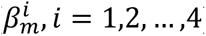) in such a way that the model generates the above-mentioned mean annual force of infection (see Table S2 for parameter values).

For the hospitalization rate, we used data published in (21), which reported the proportion of dengue cases hospitalized in different age groups during 2003 to 2017 in Singapore. During 2003 to 2006 the hospitalization rates were reported to be exceptionally high due to less strict admission criteria of dengue patients in Singapore. We therefore consider the data reported during 2007 to 2017 and calculated average age-specific hospitalization rate (see Table S5). We further assume that the rate at which secondary cases are hospitalized (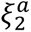) is four times higher than that of primary cases (28,29) (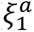), i.e., 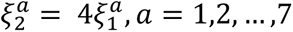.

We use vaccine efficacy estimates from the trial (18). Where they were not estimable due to insufficient hospitalization events among vaccine groups for DENV-4 in both seropositive and seronegative individuals, and for DENV-2 in seronegative individuals, we assume that efficacy estimates against hospitalization are same as those against VCD for that group (see Table S3).

### Model fitting

We fit the model to the data on age-stratified annual incidence of dengue cases per 100,000 population (Singaporean citizen and PR) during 2014 to 2020, collected from the reports on communicable disease surveillance published by Ministry of Health (MOH), Singapore (30). The data were reported for the following eight age groups: 0–4, 5–14, 15–24, 25–34, 35–44, 45–54, 55–64, and 65+ years. We used a Bayesian framework implanted using *“odin.dust”* and *“mcstate”* libraries (31) in R for model fitting and estimated eight age-specific fractions of secondary infections reported to as cases (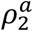 *for a* = 1,2, …, 8). We assume that these reporting rates of secondary infections are two times higher than the reporting rates of primary infections (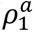 *for a* = 1,2, …, 8) i.e., 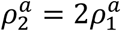 *for a* = 1,2, …, 8 (28). We used Poisson likelihood and chose uniform priors (𝒰[0,1]) for all the estimated parameters. The model fitting is presented in Fig S3. The estimated parameters values are presented in Table S4 and the trace plot of the MCMC chain is presented in Fig S4.

### Scenarios

We consider four different scenarios based on the dominant dengue serotypes during vaccine rollout. These scenarios are created by importing small number of serotype-specific infections over time so that the cumulative number of infections due to this specific serotype during the vaccine rollout results in highest contribution compared to the remaining serotypes (see Fig S5, for the underlying serotype distribution in each of the scenarios). In each of the scenarios, we target seven different targeted age-groups for vaccination: 6–16 (WHO-SAGE recommended age group for vaccination with Qdenga in high dengue endemic regions), 17–30, 31–40, 41–50, 51– 60, 61–70, and 71–80 years. We also consider three levels of vaccine coverage: low (20%), medium (50%), and high (80%), for these age groups.

To assess the differential benefits of vaccination based on baseline serostatus, we also estimated the impact separately among seropositive and seronegative individuals under each serotype-dominant scenario and targeted age group.

Although WHO does not currently recommend pre-vaccination screening for Qdenga, we consider a screening scenario in which vaccination is preceded by a serological test with 90% sensitivity and specificity. We then compare the population- and individual-level impact of this strategy against the standard Qdenga vaccination program without pre-screening.

Since clinical trial data for Qdenga does not provide conclusive evidence on its efficacy against dengue virus infection, we assumed no protection against infection in the main analysis. To explore an alternative scenario, we also evaluated the vaccination impact assuming Qdenga provides protection against infection similar to that of Dengvaxia. We used efficacy estimates reported for Dengvaxia, with 9·3% (95% CI: –35·9–38·8) for seronegative individuals and 48·1% (95% CI: 35·2–58·5) for seropositive individuals, against any serotype (32).

### Evaluation of vaccine impact

We quantify the population-level impact of vaccination as the percentage reduction in annual dengue cases and hospitalizations across the entire population over a 10-year vaccination program. For subgroup-specific analyses, such as those among seropositive individuals, seronegative individuals, among vaccinated individuals and specific age groups, we used the same metric, calculated within each respective cohort.

To evaluate the impact of vaccine, for each model run we randomly draw 100 samples from the posterior distribution of the estimated age-specific reporting rates and vaccine efficacy (VE) estimates from skew normal distribution (with shape parameter 1) using the point estimates and confidence intervals reported in the clinical trials (18), and calculate the mean and 95% range of simulations, of the relevant quantities. All the remaining parameters, including the serotype specific disease transmission rates, are kept fixed as presented in Table S2.

### Sensitivity analysis

We performed sensitivity analyses by relaxing the baseline assumption (Assumption 1) that reporting rates and hospitalization rates depend both on age and pre-exposure history. Specifically, we considered two alternative assumptions: (i) both reporting rates and hospitalization rates are age-specific but independent of pre-exposure history (Assumption 2) and (ii) reporting rates remain dependent on pre-exposure history as in the baseline model, while hospitalization rates are independent of pre-exposure history (Assumption 3).

Additionally, we explore an alternative vaccination scenario targeting broader age cohorts, which may be more practical to implement and allows vaccination of a larger number of individuals. For this analysis, we consider the targeted age groups as 6–16, 17–30, 31–60, 30+, 40+, 50+, and 60+ years.

## Results

### Population-level impact of vaccination on reported dengue cases and hospitalization

In DENV-1–4 dominant scenarios, we find that an 80%-coverage routine Qdenga program can avert up to 9% (95% range: 8%–11%), 12% (95% range: 11%–12%), 7% (95% range: 5%–9%), and 5% (95% range: –0.5%–10%) of reported dengue cases respectively (Fig 1(a)–(d)). Because we assume no vaccine efficacy against infection (and hence no indirect effects), the impact both on cases and hospitalizations scales linearly with coverage (Fig 1).

**Fig 1:**
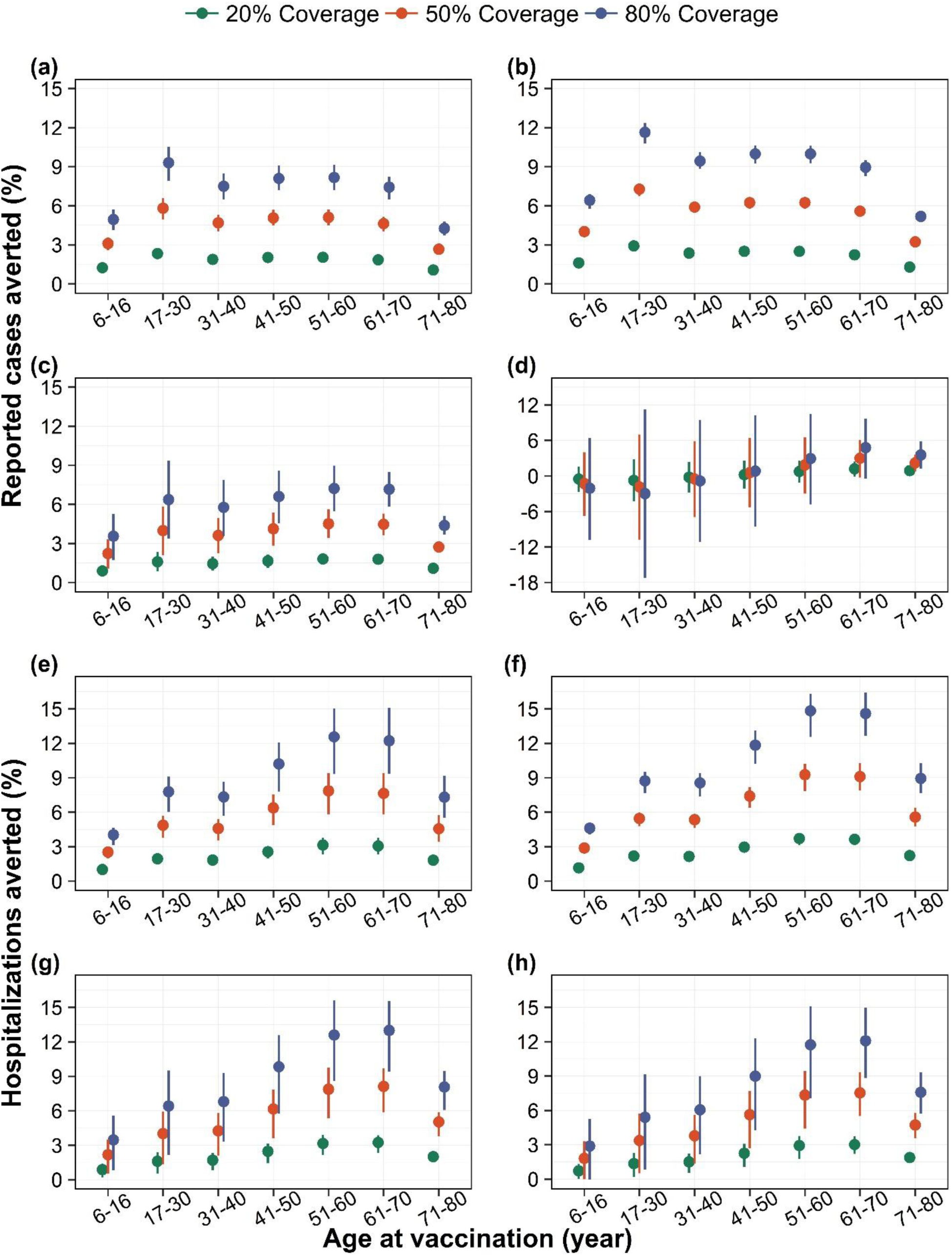
Population impact of vaccination on dengue cases and hospitalizations. Percentage of reported dengue cases averted by targeted age groups under (a) DENV-1, (b) DENV-2, (c) DENV-3, and (d) DENV-4 dominant scenarios, and percentage of hospitalizations due to dengue averted by targeted age groups under (e) DENV-1, (f) DENV-2, (g) DENV-3, and (h) DENV-4 dominant scenarios, for three different vaccine coverage levels, over a 10-year routine vaccination program with Qdenga. Solid dots represent the mean model projections, and error bars indicate the corresponding 95% range of simulations.

The targeted age group for vaccination plays a key role in determining the population-level impact on dengue cases, with the optimal age group differing across serotype-dominant scenarios (see Fig 1(a)–(d)). In the scenarios, where DENV-1 and DENV-2 are dominant, targeting those who are in the age group 17–30 years for vaccination, may result in maximum population-level impact in terms of incidence of cases averted. However, in DENV-3 and 4 dominant scenarios, the mean estimates indicate a higher impact when older age groups are targeted for vaccination.

Vaccination may avert on average 12%–15% of hospitalizations at 80% coverage across all dominant serotype scenarios (Fig 1 (e)–(h)). Like cases averted, the impact of vaccination on hospitalization depends on targeted age-groups but the optimal age group remains consistent throughout all four scenarios, estimating that targeting older age groups (i.e. 51–60 years and 61– 70 years) for vaccination can avert maximum dengue hospitalizations (Fig 1 (e)–(h)). By contrast, following the WHO-SAGE recommended strategy of vaccinating children aged 6–16 years at 80% coverage would avert around 6% of reported cases and 5% of hospitalizations across all four serotype-dominant scenarios (Fig 1).

To assess the robustness of our findings, we evaluated the impact of relaxing the baseline assumptions regarding the dependence of reporting rates and hospitalization rates on pre-exposure history. While the overall qualitative patterns of vaccine impact across serotype-dominant scenarios and age groups remained consistent, the predicted vaccine impacts under DENV-3 and 4 dominant scenarios were generally lower and accompanied by greater uncertainty when the exposure dependence of reporting and/or hospitalization rates was relaxed (i.e., Assumption 2 and Assumption 3) compared to the baseline assumption (Assumption 1) (see Fig S6 and Fig S7).

We examined the impact of vaccination both within targeted age groups and in adjacent age groups that individuals aged into over time. Under DENV-1 and DENV-2 dominant scenarios, the impact on both cases and hospitalizations remained stable across targeted age groups. However, under DENV-3 and DENV-4 dominant scenarios, the impact increased monotonically with older targeted age groups. Across all four serotype-dominant scenarios, we observed an overall increasing trend in impact as older age groups were vaccinated, although this increase was not strictly monotonic (see Fig S8 and S9).

When broader age groups were considered, targeting individuals aged 30 years and above, the age group with the highest number of cumulative vaccinees over a 10-year routine program (see Fig S12), consistently yielded the maximum reductions in both dengue cases and hospitalizations across all four serotype-dominant scenarios (see Fig S10 and S11).

### Impact of vaccination according to baseline serostatus

We further estimate the impact of vaccine among seropositive and seronegative individual separately. In DENV-1 and DENV-2 dominant scenarios, both seronegative and seropositive populations experience a positive impact both in terms of cases and hospitalization reduction (Fig 2 (a), (b), (e), and (f)). In DENV-3 and DENV-4 dominant scenarios, the vaccination impact among seropositive individuals remains consistently positive. However, among seronegative individuals, the mean impact can be relatively low (as in the DENV-3 scenario for cases) or even negative (as in the DENV-4 for cases and DENV-3 and 4 scenarios for hospitalization), with greater uncertainty (Fig 2 (c), (d), (g), and (h)).

**Fig 2:**
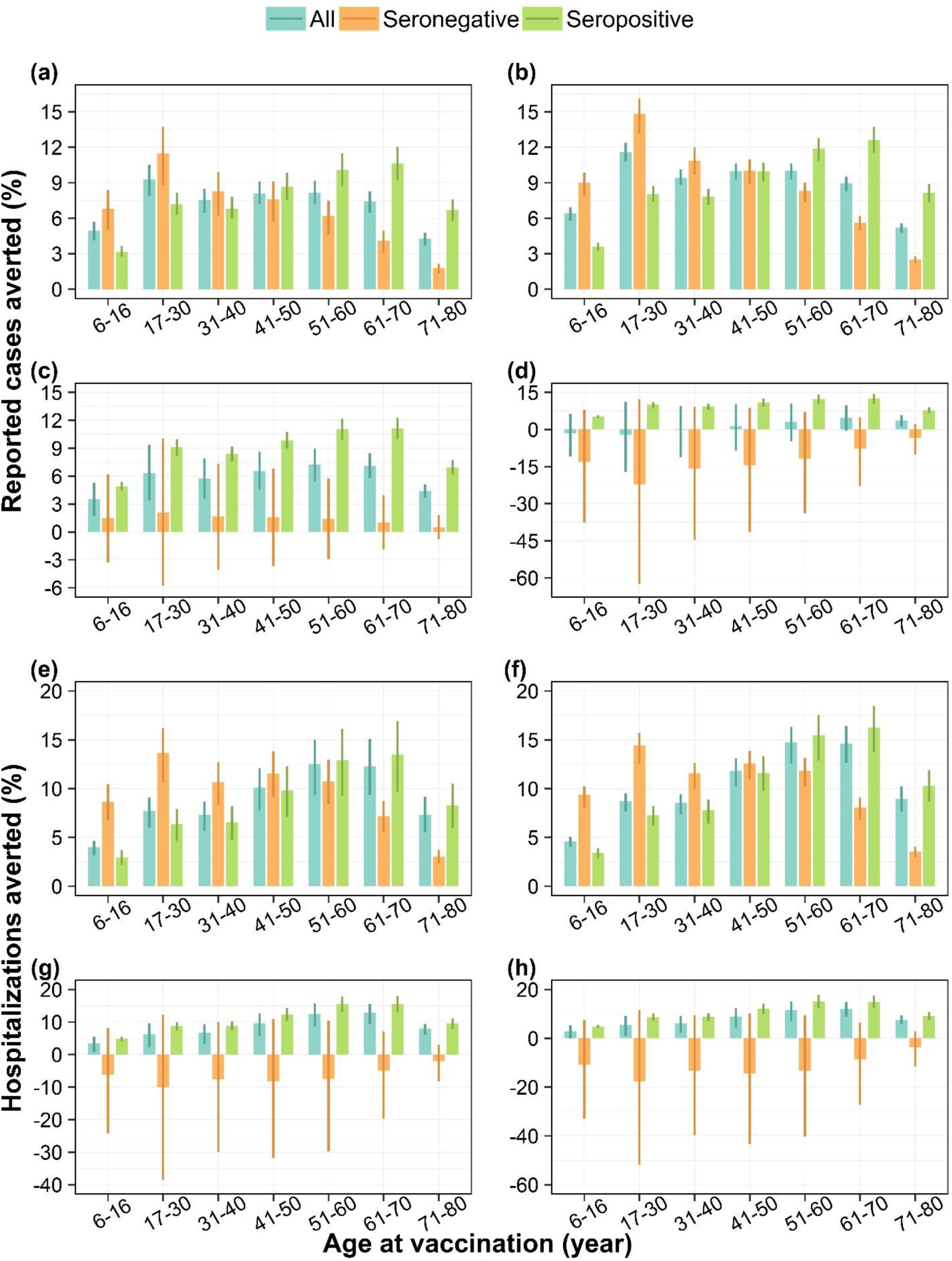
Impact of vaccination on dengue cases and hospitalizations stratified by baseline serostatus. Percentage of reported dengue cases averted by targeted age groups under (a) DENV-1, (b) DENV-2, (c) DENV-3, and (d) DENV-4 dominant scenarios, and percentage of hospitalizations due to dengue averted by targeted age groups under (e) DENV-1, (f) DENV-2, (g) DENV-3, and (h) DENV-4 dominant scenarios, for the whole population, seropositive individuals only, and seronegative individuals only, assuming 80% vaccine coverage, over a 10-year routine vaccination program with Qdenga. Bars represent mean model estimates, and error bars indicate the corresponding 95% range of simulations.

### Impact of pre-screening

Routine vaccination program with a pre-screening strategy reduces the overall population impact across all serotype-dominant scenarios (Fig 3(a)–(d) and Fig 4(a)–(d)). However, we find that among vaccinated individuals pre-screening does not improve the impact under DENV-1 and 2 dominant scenarios (Fig 3(e)–(f) and Fig 4(e)–(f)) but provides an additional benefit under DENV-3 and 4 dominant scenarios (Fig 3(g)–(h) and Fig 4(g)–(h)).

**Fig 3:**
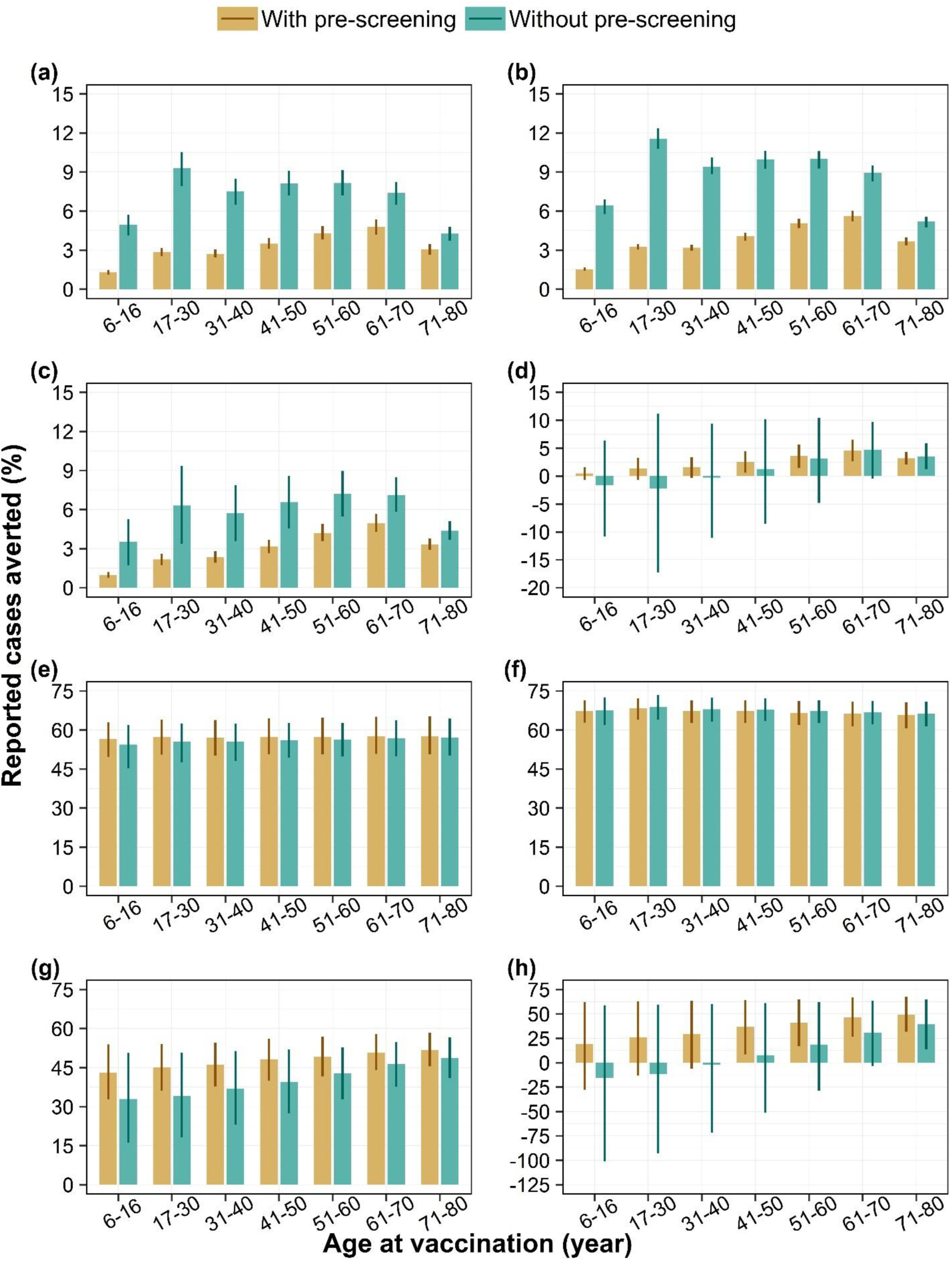
Impact of vaccination with and without pre-screening on dengue cases. Percentage of reported dengue cases averted by targeted age groups over a 10-year routine vaccination program with 80% coverage, among the whole population (i.e., population impact) under (a) DENV-1, DENV-2, (c) DENV-3, and (d) DENV-4 dominant scenarios and among vaccinated individuals only (i.e., individual impact) under (e) DENV-1, (f) DENV-2, (g) DENV-3, and (h) DENV-4 dominant scenarios, with diagnostic pre-screening versus no screening. Bars represent mean model estimates, and error bars indicate the corresponding 95% range of simulations.

**Fig 4:**
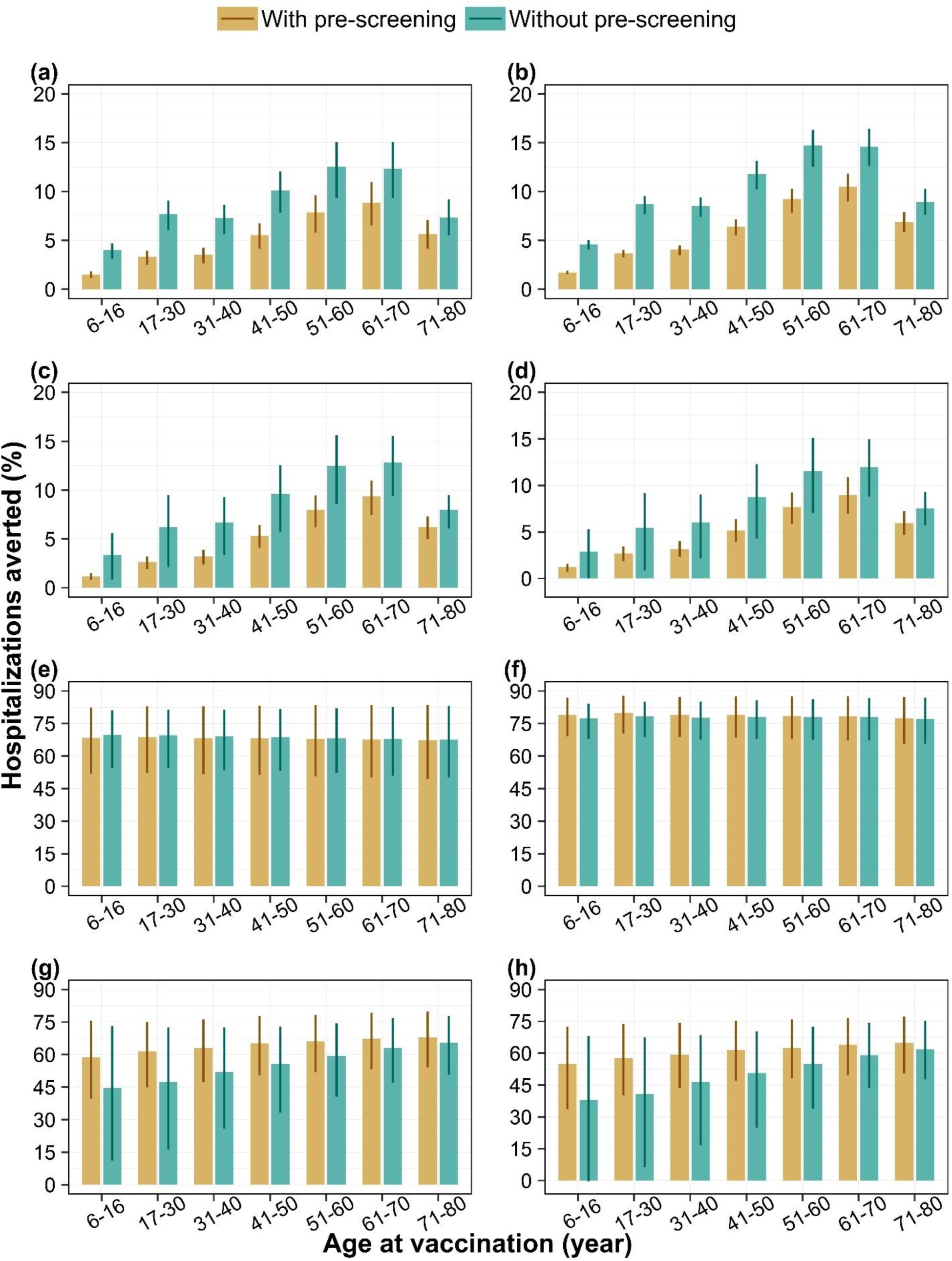
Impact of vaccination with and without pre-screening on dengue hospitalizations. Percentage of reported dengue hospitalizations averted by targeted age groups over a 10-year routine vaccination program with 80% coverage, among the whole population (i.e., population impact) under (a) DENV-1, (b) DENV-2, (c) DENV-3, and (d) DENV-4 dominant scenarios and among vaccinated individuals only (i.e., individual impact) under (e) DENV-1, (f) DENV-2, (g) DENV-3, and (h) DENV-4 dominant scenarios, with diagnostic pre-screening versus no screening. Bars represent mean model estimates, and error bars indicate the corresponding 95% range of simulations.

### Impact with efficacy against infection

Irrespective of serotype-dominant scenarios, the mean impact on both cases and hospitalizations is always higher in the presence of efficacy of vaccine against infection compared to its absence (Fig 5(a)–(h)). Across serotype-dominant scenarios, with this additional efficacy against infection, vaccination can avert up to 47% (95% range: 18%–60%) of cases and 52% (95% range: 23%–64%) hospitalizations. In the DENV-1 and DENV-2 dominant scenarios, including efficacy against infection shifts the age group yielding the greatest population-level case reduction to an older group than in the scenario without efficacy against infection.

**Fig 5:**
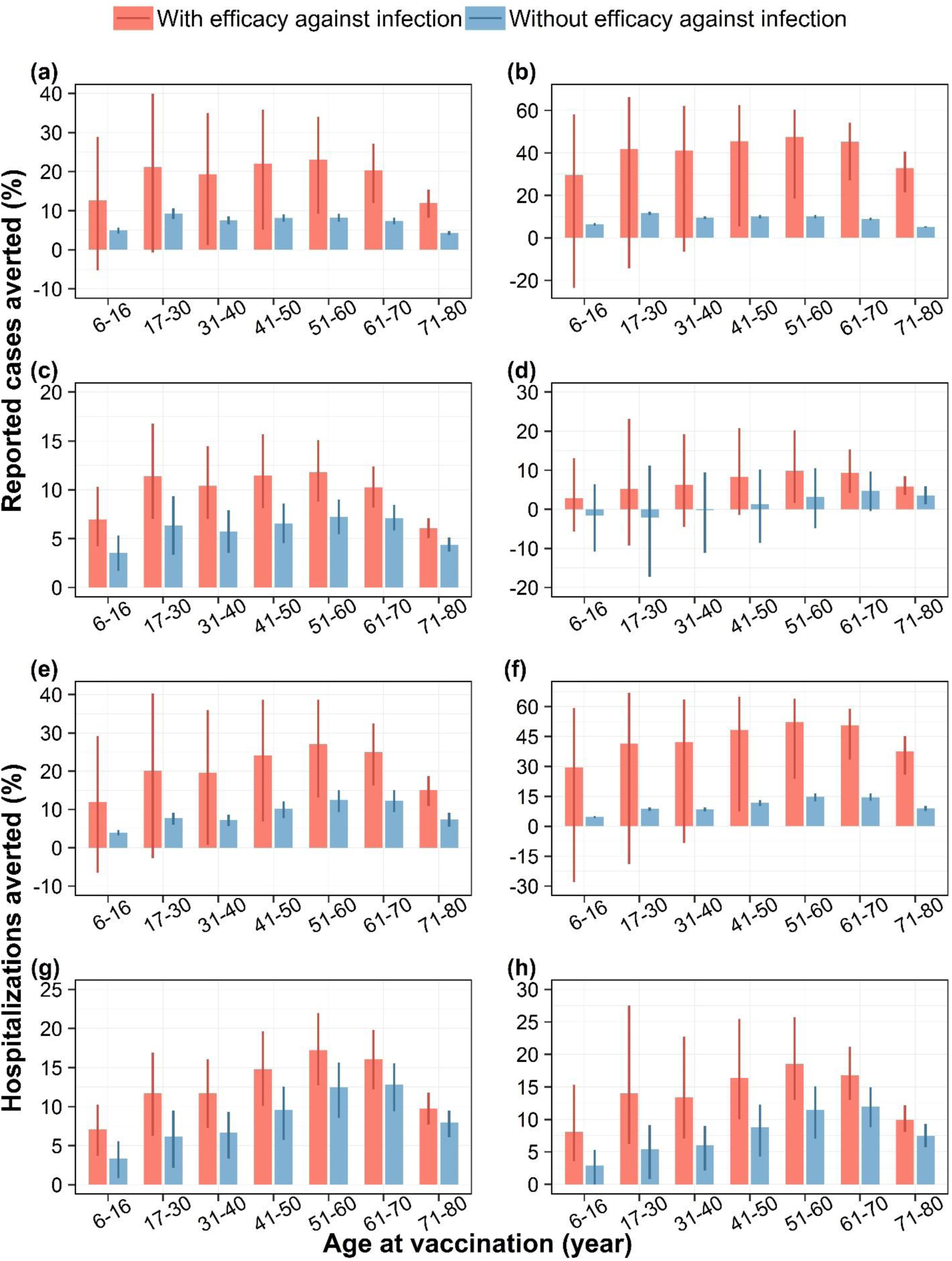
Impact of vaccination on dengue cases and hospitalizations, with and without efficacy against infection. Percentage of reported dengue cases averted by targeted age groups under (a) DENV-1, (b) DENV-2, (c) DENV-3, and (d) DENV-4 dominant scenarios, and percentage of hospitalizations due to dengue averted by targeted age groups under (e) DENV-1, (f) DENV-2, (g) DENV-3, and (h) DENV-4 dominant scenarios, assuming the vaccine has efficacy against infection with any serotype similar to that of Dengvaxia, over a 10-year routine vaccination program with 80% vaccine coverage. Bars represent mean model estimates, and error bars indicate the corresponding 95% range of simulations.

## Discussion

Based on an age-stratified, multi-serotype transmission modelling framework, our analysis suggests that routine dengue vaccination with Qdenga (TAK-003) in Singapore could avert up to 12% of dengue cases and 15% of hospitalizations over a 10-year period at 80% coverage, across dominant serotype scenarios. The impact of vaccination varies by targeted age groups, with the greatest reduction achieved by vaccinating individuals aged 17–30 years for cases and 51–70 years for hospitalizations.

To explore the potential impact of vaccination under differing epidemiological conditions, we constructed four serotype-dominant scenarios, each characterized by the dominance of a specific dengue serotype. As these scenarios were created under different assumptions about the introduction and combination of serotypes, the projected vaccine impacts are not intended to be directly compared across scenarios, but rather to illustrate how performance of vaccination may vary depending on the prevailing serotype. Based on these scenarios, our model, consistent with clinical trial observations, indicated that the public health benefit of Qdenga would be most pronounced in scenarios dominated by DENV-1 and DENV-2, the serotypes against which the vaccine had demonstrated higher efficacy. In these scenarios, routine vaccination programs with higher vaccine coverage (i.e., 80%) can lead to substantial reductions in dengue-related morbidity, including up to 12% of reported cases and 15% of hospitalizations in Singapore. In contrast, the projected impact is lower and more uncertain in DENV-3 and DENV-4 dominant scenarios.

The vaccine impact estimates were evaluated based on the assumption, supported by epidemiological evidence that secondary infections are more likely to be symptomatic and lead to hospitalization compared to primary infections. This assumption, which has also been adopted in prior modelling studies (28,29), is epidemiologically plausible given the increased severity typically associated with secondary infections. Our sensitivity analysis indicated that while the overall qualitative patterns of vaccine impact are robust, relaxing the exposure dependence of reporting and hospitalization rates leads to lower projected impact particularly in DENV-3 and DENV-4 dominant scenarios. This may be attributed to the fact that, under the alternative assumptions, seropositive and seronegative individuals are equally likely to be reported or hospitalized. Given the relatively low population immunity against dengue in Singapore, a larger proportion of reported cases or hospitalizations under these assumptions are likely to be seronegative vaccinees, for whom the vaccine did not demonstrate protective efficacy, particularly against DENV-3 and DENV-4. However, availability of more granular epidemiological data in future, particularly with classification based on infection history, would help to improve parameterization and reduce uncertainty in model-based projection.

Our results underscore the importance of tailoring vaccination strategies to local epidemiological conditions. In Singapore, routine vaccination targeting the WHO-SAGE recommended 6–16-year age group had limited impact compared to strategies focusing on older age groups. In Singapore, the age distribution of dengue infection has shifted toward older population due to low transmission intensity. Recent surveillance data indicate that dengue cases are increasingly concentrated among individuals aged 15-35 years, while hospitalizations are more common among those older than 45 years. Consequently, our model predicted that prioritizing older age groups, which aligns with the current burden of dengue infection, would result in greater overall vaccine impact.

We also considered an alternative strategy of targeting broader age groups for vaccination, which may be more practical to implement. In Singapore’s low-transmission setting, this approach would still include individuals with only a single prior infection, who are likely to benefit from vaccination. This contrasts with high-transmission settings, where individuals beyond the recommended 6–16-year age group are more likely to have experienced multiple infections and thus derive limited additional benefit. This strategy also allows for the inclusion of both younger adults at higher risk of dengue cases and older adults at higher risk of hospitalization, aligning with the current distribution of disease risk in Singapore.

The safety and efficacy of Qdenga in older age groups remain largely unknown, as clinical data are limited beyond individuals aged 4–16 years. Our projections on vaccine impact assumed that vaccine efficacy in older age groups is comparable to that observed in 4–16 years old individuals, based on an immunobridging study (22) indicating similar immunogenicity. However, the WHO recommended the use of Qdenga up to the age of 60 years (12), which supports the relevance of evaluating broader age-based vaccination strategies. For policy decisions, especially in settings considering expanded age-based vaccination strategies, this highlights the importance of post-licensure surveillance or targeted effectiveness studies to validate these assumptions. While our model-based analysis explored the potential benefits of broader age-targeted vaccination, these findings should be interpreted with the understanding that estimates may need to be updated as further evidence becomes available.

Our baseline serostatus stratified impact analysis reinforces the potential benefits of vaccination among seropositive individuals, who consistently show reductions in both cases and hospitalizations across all serotype-dominant scenarios. However, the risk-benefit balance is less favorable for seronegative individuals in DENV-3 and DENV-4 dominant scenarios, where the vaccination may offer limited or even negative impact. This aligns with our finding that the optimal age group for vaccination shifts toward older populations under DENV-3 and DENV-4 scenarios—likely reflecting both the lower efficacy among seronegative individuals for these serotypes and the higher baseline seroprevalence in older age groups in Singapore.

We explored the potential benefit of pre-screening as a strategy to reduce the proportion of seronegative individuals in the vaccinated cohort, aiming to enhance the overall impact. Our model predicted that implementing such strategy could reduce the population impact on both cases and hospitalizations, possibly due to Singapore’s relatively low proportion of seropositive individuals means that fewer people would qualify for vaccination under a *screen-and-vaccinate* approach. This also aligns with the results reported in a recent modelling study (33), which found that pre-vaccination screening significantly reduced the population-level impact across transmission settings, most notably in low-transmission setting. Our findings also suggest that pre-screening does not provide additional benefits in averting cases and hospitalizations among vaccinated individuals in DENV-1 and DENV-2 dominant scenarios, as vaccine showed similar level of protection among both seropositive and seronegative individuals. In contrast, for DENV-3 and DENV-4 dominant scenarios, pre-screening was found to offer an additional benefit in reducing both cases and hospitalizations among vaccinated. This is because there is a significant difference in vaccine efficacy estimates between seropositive and seronegative individuals, with negative efficacy and wider uncertainty observed in the seronegative group. Given this uncertainty, the WHO has planned a post-authorization effectiveness study in Southeast Asia to further assess the impact of Qdenga on severe dengue and hospitalizations due to DENV-3 and DENV-4 (WHO, 2023). We will revisit our estimates of vaccine impact once the results from this study become available.

While our main impact analysis assumes no vaccine-induced protection against infection, we explored a scenario where Qdenga provides protection akin to Dengvaxia against infection due to any serotype. As expected, the inclusion of such efficacy leads to higher impact estimates across all scenarios, suggesting that even modest protection against infection could yield meaningful indirect benefits through reduced transmission. However, given the lack of conclusive evidence for Qdenga’s effect on infection risk, these results should be interpreted cautiously.

Although our study uses a comprehensive modeling framework that explicitly accounts for age structure, serotype-specific transmission, and baseline serostatus, along with corresponding variations in vaccine efficacy to estimate the vaccine impact more accurately, it has several limitations. We assumed that vaccine efficacy remains constant over time, based on the cumulative protection observed 54 months after the second dose. While this implicitly incorporates some degree of waning, further decline in protection may occur beyond this time frame. In practice, if vaccine-induced immunity decreases substantially, booster doses would likely be introduced to sustain protection. The potential role of booster doses in maintaining long-term vaccine efficacy is currently being evaluated in ongoing studies, and their implementation could help sustain the protection over time. The impact estimates presented here reflect the potential benefit under a maintained efficacy scenario. However, the associated uncertainty in longer-term projections should be considered when interpreting the results.

This study focuses solely on evaluating the impact of vaccination, it does account for routine dengue control interventions in Singapore implicitly as we use estimates of the force of infection in Singapore during which routine vector control was in place. We do not include any future impact of the Wolbachia mosquito release program. However, since our model explicitly incorporated the mosquito population, it provides a flexible framework and will be extended in future work to evaluate the combined impact of vaccination and mosquito control programs.

Our study quantified the public health benefits of vaccination in terms of averted cases and hospitalizations. In future work we will perform a cost-effectiveness analysis which will provide a more complete picture of the impact by incorporating health economic metrics such as disability-adjusted life years (DALYs) or quality-adjusted life years (QALYs). Unlike the previous study by (10) that investigated cost effectiveness of future vaccination in Singapore with a static model, our dynamic transmission model-based framework can serve as a robust foundation for such an evaluation. Moreover, our framework is easily adaptable to other dengue-endemic regions and can be applied to assess the impact of upcoming dengue vaccines, such as Butantan-DV.

Our analysis considered only the resident population of Singapore (citizens and permanent residents), excluding foreign workers who make up nearly one-third of the total population. Routine dengue surveillance is limited in this group, particularly among those employed in high-risk settings such as construction sites, marine yards, and processing sectors, which often present favorable breeding conditions for mosquitoes. Given their elevated infection risk and limited surveillance data, we are currently extending our modelling framework to include this population and evaluate vaccination strategies tailored to this group

In conclusion, our analysis suggests that targeting adult age groups for routine dengue vaccination with Qdenga in Singapore may yield greater public health impact, consistent with the current age distribution of dengue cases and hospitalizations. Our modelling work highlights the need for careful evaluation of the local epidemiological and demographic context and provides key insights into the roles of serotype distribution, baseline serostatus, age-specific targeting, and the vaccine’s complex efficacy profile in designing effective dengue vaccination strategies.

## Data Availability

No new primary data was collected for this modelling study. All the code for reproducing the results can be found at https://github.com/ID-Modelling-Lab/singapore_dengue_vaccination.

## Acknowledgements

This research is supported by the Singapore Ministry of Health’s National Medical Research Council under its Open Fund – Individual Research Grant (MOH-001118-00). The funder has no specific role in the conceptualization, design, data collection, analysis, decision to publish, or preparation of the manuscript.

## Author contributions

AS did the model development, formal analysis, conceptualized the methodology, created the software, wrote the original draft and revised and edited the manuscript. TLT did the model development and reviewed the manuscript. YSL did the model development and reviewed the manuscript. HEC conceptualized the study, did the model development, acquired funding, conceptualized the methodology, investigated, supervised, and reviewed and edited the manuscript. All authors read and approved the final version of the manuscript.

## Ethical approval

No human subjects were involved in this work and therefore ethical approval was not required.

