## Supplementary Information for "Adult dengue vaccination in Singapore: A modelling study to inform policy"

### 1. Mathematical equations of the transmission model

$$\frac{dS^a}{dt} = \lambda^h - \left(\sum_{i=1}^{4} \lambda_i^m + \mu^h\right) S^a + a_+ S^{a-1} - a_- S^a - v_{S^a},$$

$$\frac{dI_i^a}{dt} = \lambda_i^m S^a - (\gamma_1 + \mu^h) I_i^a + a_+ I_i^{a-1} - a_- I_i^a,$$

$$\frac{dC_i^a}{dt} = \gamma_1 I_i^a - (\alpha + \mu^h) C_i^a + a_+ C_i^a - a_- C_i^a - v_{C_i^a},$$

$$\frac{dS_i^a}{dt} = \alpha C_i^a - \left(\sum_{j \neq i} \lambda_j^m + \mu^h\right) S_i^a + a_+ S_i^{a-1} - a_- S_i^a - v_{S_i^a},$$

$$\frac{dI_{ij}^a}{dt} = \lambda_j^m S_i^a - (\gamma_2 + \mu^h) I_{ij}^a + a_+ I_{ij}^{a-1} - a_- I_{ij}^a,$$

$$\frac{dR^a}{dt} = \gamma_2 \sum_{i,j} I_{ij}^a - \mu^h R^a + a_+ R^{a-1} - a_- R^a - v_{R^a}, \tag{S1}$$

$$\frac{dS^m}{dt} = \Lambda^m - \left(\sum_{i=1}^{4} \lambda_i^h + \mu^m\right) S^m,$$

$$\frac{dE_i^m}{dt} = \lambda_i^h S^m - (\sigma^m + \mu^m) E_i^m,$$

$$\frac{dI_i^m}{dt} = \sigma^m E_i^m - \mu^m I_i^m,$$

$$\frac{dS_v^a}{dt} = v_{S^a} - \left(1 - \epsilon^{inf-}\right) \left(\sum_{i=1}^{4} \lambda_i^m + \mu^h\right) S_v^a + a_+ S_v^{a-1} - a_- S_v^a,$$

$$\frac{dI_{vi}^a}{dt} = \left(1 - \epsilon^{inf-}\right) \lambda_i^m S_v^a - (\gamma_1 + \mu^h) I_{vi}^a + a_+ I_{vi}^{a-1} - a_- I_{vi}^a,$$

$$\frac{dC_{vi}^{a}}{dt} = \nu_{C_i^a} + \gamma_1 I_{vi}^{a} - (\alpha + \mu^h) C_{vi}^{a} + a_+ C_{vi}^{a-1} - a_- C_{vi}^{a},$$

$$\frac{dS_{vi}^{a}}{dt} = \nu_{S_i^a} + \alpha C_{vi}^{a} - \left(1 - \epsilon_s^{inf+}\right)\left(\sum_{j\neq i} \lambda_j^m + \mu^h\right) S_{vi}^{a} + a_+ S_{vi}^{a-1} - a_- S_{vi}^{a},$$

$$\frac{dI_{vij}^{a}}{dt} = \left(1 - \epsilon^{inf+}\right)\lambda_j^m S_{vi}^{a} - (\gamma_2 + \mu^h) I_{vij}^{a} + a_+ I_{vij}^{a-1} - a_- I_{vij}^{a},$$

$$\frac{R_v^a}{dt} = \nu_{R^a} + \gamma_2 \sum_{i,j} I_{vij}^{a} - \mu^h R_v^a + a_+ R_v^{a-1} - a_- R_v^a.$$

The force of infection on human due to serotype *i,* is given by

$$\lambda_i^m = \frac{b\beta_i^m I_i^m}{N} \qquad (S2)$$

The force of infection on mosquitoes from human infected with serotype *i*, is given by

$$\lambda_i^h = \frac{b\beta_i^h \left(\sum_a I_i^a + \sum_{a,j(j\neq i)} I_{ji}^a + \sum_a I_{vi}^a + \sum_{a,j(j\neq i)} I_{vji}^a\right)}{N}. \qquad (S3)$$

The incidence of infection of age *a* at time *t* due to serotype *i* is given by,

$$Inf_i^a(t) = \lambda_i^m S^a + \left(1 - \epsilon^{inf-}\right)\lambda_i^m S_v^a + \sum_{j(\neq i)} \lambda_j^m S_i^a + \sum_{j(\neq i)} \left(1 - \epsilon^{inf+}\right)\lambda_j^m S_{vi}^a \qquad (S4)$$

The incidence of reported cases of age *a* at time *t* due to serotype *i* is given by,

$$Rep_i^a(t) = \rho_1^a \lambda_i^m S^a + \rho_2^a \sum_{j(\neq i)} \lambda_j^m S_i^a + \rho_1^a \sum_{s} \left(1 - \epsilon_i^{vcd-|inf-}\right)\left(1 - \epsilon^{inf-}\right)\lambda_i^m S_v^a +$$
$$\rho_2^a \sum_{j(\neq i)} \left(1 - \epsilon_i^{vcd+|inf+}\right)\left(1 - \epsilon^{inf+}\right)\lambda_j^m S_{vi}^a \qquad (S5)$$

The incidence of hospitalization of age *a* at time *t* due to serotype *i* is given by,

$$Hosp_i^a(t) = \xi_1 \rho_1^a \lambda_i^m S^a + \xi_2 \rho_2^a \sum_{j(\neq i)} \lambda_j^m S_i^a +$$
$$\xi_1 \rho_1^a \sum_s \left(1 - \epsilon_i^{hosp-|vcd-}\right)\left(1 - \epsilon_i^{vcd-|inf-}\right)\left(1 - \epsilon^{inf-}\right) \lambda_i^m S_v^a +$$
$$\xi_2 \rho_2^a \sum_{j(\neq i)} \left(1 - \epsilon_i^{hosp+|vcd+}\right)\left(1 - \epsilon_i^{vcd+|inf+}\right)\left(1 - \epsilon^{inf+}\right) \lambda_j^m S_{vi}^a \qquad (S6)$$

| Symbol | Description |
|---|---|
| $S^a(t)$ | Number of people of age *a* at time *t*, who are susceptible to infection from any serotype |
| $I_i^a(t)$ | Number of people of age *a* at time *t*, who are infected with serotype *i* (primary infection) |
| $C_i^a(t)$ | Number of age *a* at time *t* who are immune to infection from serotype *i*, and temporarily protected against heterologous infection |
| $S_i^a(t)$ | Number of people of age a at time *t* who are immune to infection from serotype *i* but remain susceptible to infection from other serotypes |
| $I_{ij}^a(t)$ | Number of people of age a at time *t* who are immune to infection from serotype *i,* but infected with serotype *j* ($\neq i$) (secondary infection) |
| $R^a(t)$ | Number of people of age a at time t who are recovered from secondary infection |
| $S_v^a(t)$ | Number of vaccinated people of age *a,* at time *t*, who are susceptible to infection from any serotype |

| | |
|---|---|
| $I_{vi}^{a}(t)$ | Number of vaccinated people of age *a,* at time *t*, who are infected with serotype *i (primary infection)* |
| $C_{vi}^{a}(t)$ | Number of vaccinated people of age *a* at time *t* who are immune to infection from serotype $i$, and temporarily protected against heterologous infection |
| $S_{vi}^{a}(t)$ | Number of vaccinated people of age *a* at time *t* who are immune to infection from serotype *i* but remain susceptible to infection from other serotypes |
| $I_{vij}^{a}(t)$ | Number of vaccinated people of age *a* at time *t* who are immune to infection from serotype $i,$ but infected with serotype *j* $(\neq i)$ (secondary infection) |
| $R_{v}^{a}(t)$ | Number of vaccinated people of age *a* at time *t* who are recovered from secondary infection |
| $N(t)$ | Total human population at time *t* |
| $S^{m}(t)$ | Number of uninfected adult female mosquitoes at time *t* |
| $E_{i}^{m}(t)$ | Number of adult female mosquitoes at time *t* in incubation period infected with serotype *i* |
| $I_{i}^{m}(t)$ | Number of infectious adult female mosquitoes infected with serotype *i* |

**Table S1:** The description of the state variables used in the transmission model.

| Parameters | Description | Value | Source |
|---|---|---|---|
| $\Lambda^h$ | Human recruitment rate | Birth rate×Total population (Time dependent) | (1,2) |
| $\mu^h$ | Mortality rate of human | Time dependent | (1) |
| $\rho_2^a$ | Fraction of secondary infection in age *a* to be reported | See Table S6 | Estimated from age-stratified annual incidence of reported dengue cases per 100,000 population from 2014-2020 |
| $\rho_1^a$ | Fraction of primary infection in age *a* to be reported | $\frac{1}{2}\rho_2^a$ | (3,4) |
| $\xi_2^a$ | Fraction of reported cases from secondary infection in age *a* | Table S5 | (5) |

| | | | |
|---|---|---|---|
| | requires hospitalization | | |
| $\xi_1^a$ | Fraction of reported cases from primary infection in age $a$ requires hospitalization | $\frac{1}{4}\xi_2$ | (3,4) |
| $\frac{1}{\gamma}$ | Infectious period of human | 4 days | (6) |
| $\frac{1}{\alpha}$ | Duration of heterologous protection from infection | 1 year | (7) |
| $b$ | Mosquito biting rate | 15 | Assumed |
| $\beta_i^h$ | Per bite probability of transmission from infected human to mosquitoes | 0.2 | Assumed |
| $\beta_i^m$ | Per bite probability of transmission from | 0.166; 0.176; 0.152; 0.144 | Calibrated to annual dengue FOI estimates |

| | | | |
|---|---|---|---|
| | infected mosquitoes with *i*th serotype to human | | |
| $\frac{1}{\sigma^m}$ | Mean extrinsic incubation period | 10 days | (8,9) |
| $\Lambda^m$ | Recruitment rate of mosquito | $\mu^m$ ×Total mosquito pop. | |
| $\mu^m$ | Adult mosquito mortality rate | 0.1 per day | (3,10) |
| $a_+$ | Rate at which individuals enter age group *a* from age group *a-1* | $\frac{1}{365}$ per day for *a = 2, 3,…,91,* and 0 for *a =1* | |
| $a_-$ | Rate at which individuals leave age group *a* and enters age group *a+1* | $\frac{1}{365}$ per day for *a = 1,3,…,90,* and 0 for *a =91* | |
| $v_x$ | Vaccination rate in compartments $x(= S^a, C_i^a, S_i^a, R^a)$ | These rates have been adjusted in such a way that coverage (given) in a targeted age group is maintained. We assumed the coverage to be 20%, 50% and 80% | |

| | | | |
|---|---|---|---|
| $\epsilon^{inf+}$ | Efficacy of vaccine against infection due to any serotype, among baseline seropositive individuals | 9·3% (95% CI: –35·9–38·8) | (11). |
| $\epsilon^{inf-}$ | Efficacy of vaccine against infection due to any serotype, among baseline seronegative individuals | 48·1% (95% CI: 35·2–58·5) | (11) |
| $\epsilon_i^{vcd+}$ | Efficacy of vaccine against VCD due to serotype *i*, among baseline seropositive individuals | See Table S3 | |
| $\epsilon_i^{vcd-}$ | Efficacy of vaccine against VCD due to serotype *i*, among | Table S3 | |

| | | |
|---|---|---|
| | baseline seronegative individuals | |
| $\epsilon_i^{hosp+}$ | Efficacy of vaccine against hospitalization due to serotype *i*, among baseline seropositive individuals | Table S3 |
| $\epsilon_i^{hosp-}$ | Efficacy of vaccine against hospitalization due to serotype *i*, among baseline seronegative individuals | Table S3 |
| $\epsilon_i^{vcd+\|inf+}$ | Efficacy of vaccine against VCD given infection due to serotype i, among baseline seropositive individuals | $\frac{\epsilon_i^{vcd+} - \epsilon^{inf+}}{1 - \epsilon^{inf+}}$ |

| | | |
|---|---|---|
| $\epsilon_i^{vcd-\|inf-}$ | Efficacy of vaccine against VCD given infection due to serotype $i$, among baseline seronegative individuals | $\frac{\epsilon_i^{vcd-} - \epsilon^{inf-}}{1 - \epsilon^{inf-}}$ |
| $\epsilon_i^{hosp+\|vcd+}$ | Efficacy of vaccine against hospitalization given VCD due to serotype $i$, among baseline seropositive individuals | $\frac{\epsilon_i^{hosp+} - \epsilon_i^{vcd+}}{1 - \epsilon_i^{vcd+}}$ |
| $\epsilon_i^{hosp-\|vcd-}$ | Efficacy of vaccine against hospitalization given VCD due to serotype $i$, among baseline seronegative individuals | $\frac{\epsilon_i^{hosp-} - \epsilon_i^{vcd-}}{1 - \epsilon_i^{vcd-}}$ |

**Table S2:** Description of the model parameters used in the transmission model.

| Vaccine efficacy against | Serotype | Seropositive | Seronegative |
|---|---|---|---|
| VCD | DENV-1 | 56·1 (44·6–65·2) | 45·4 (26·1–59·7) |
| | DENV-2 | 80·4 (73·1–85·7) | 88·1 (78·6–93·3) |
| | DENV-3 | 52·3 (36·7–64·0) | –15·5 (–108·2–35·9) |
| | DENV-4 | 70·6 (39·9–85·6) | –105·6 (–628·7–42·0) |
| Hospitalization | DENV-1 | 66·8 (37·4–82·3 | 78·4 (43·9–91·7) |
| | DENV-2 | 95·8 (89·6–98·3) | **88·1 (78·6–93·3)** |
| | DENV-3 | 74·0 (38·6–89·0) | –87·9 (–573·4–47·6) |

| | | | |
|---|---|---|---|
| | DENV-4 | **70·6 (39·9–85·6)** | **–105·6 (–628·7–42·0)** |

**Table S3:** Vaccine efficacy estimates of Qdenga in the safety set, approximately 57 months after the first dose, against virologically confirmed dengue (VCD) and hospitalization, stratified by baseline serostatus, serotype**.** The efficacy estimates against hospitalizations due to DENV-2 among seronegative individuals, due to DENV-4 among both seropositive and seronegative individuals are assumed to be same (highlighted in bold) as the corresponding efficacy estimates against VCD, as they were not estimable during the trials.

| **Parameter** | **Estimated value**<br>**Median (95% Credible Interval)** |
|---|---|
| $\rho_2^{0-4y}$ | 0·086 (0·077–0·095) |
| $\rho_2^{5-14y}$ | 0·286 (0·271–0·303) |
| $\rho_2^{15-24y}$ | 0·468 (0·448–0·489) |
| $\rho_2^{25-34y}$ | 0·429 (0·410–0·448) |
| $\rho_2^{35-44y}$ | 0·404 (0·386–0·423) |
| $\rho_2^{45-54y}$ | 0·415 (0·396–0·433) |
| $\rho_2^{55-64y}$ | 0·421 (0·401–0·441) |
| $\rho_2^{65+y}$ | 0·555 (0·527–0·583) |

**Table S4:** Estimated value of age-specific fraction of secondary infection to be reported as dengue cases.

| Parameter | Average hospitalization rate |
|---|---|
| $\xi_2^{0-14y}$ | 0·38 |
| $\xi_2^{15-24y}$ | 0·36 |
| $\xi_2^{25-34y}$ | 0·33 |
| $\xi_2^{35-44y}$ | 0·39 |
| $\xi_2^{45-54y}$ | 0·46 |
| $\xi_2^{55-64y}$ | 0·52 |
| $\xi_2^{65+y}$ | 0·49 |

**Table S5:** Average age-specific hospitalization rate. These data has been calculated from the data during 2007 to 2017, reported in *(5)*.

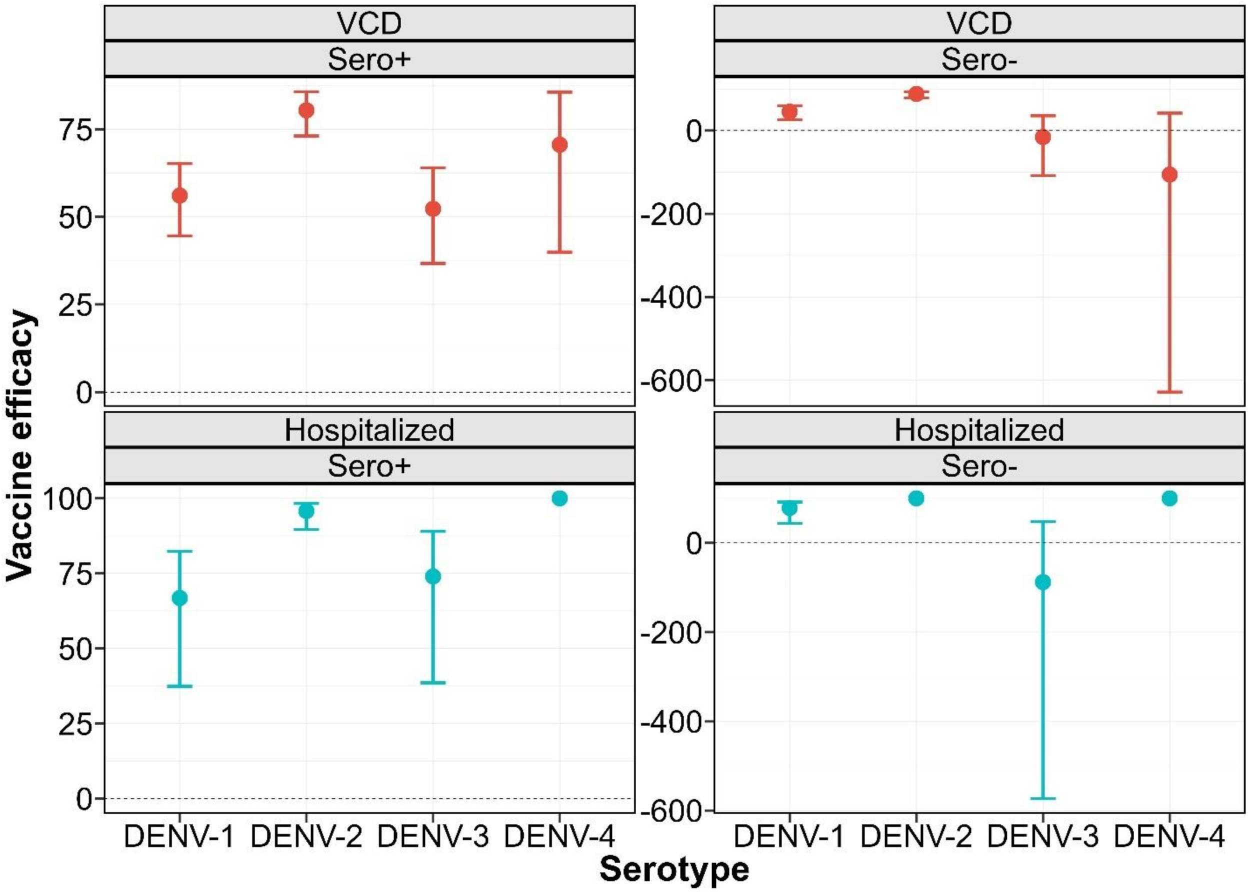

**Fig S1:** The serotype (DENV-1-4) and baseline serostatus (seropositive (Sero+), seronegative (Sero-)) stratified vaccine efficacy estimates of Qdenga against VCD and hospitalization approximately 57 months after the first dose of the vaccine. The dots represent the point estimates, and the error bar represents the 95% confidence interval. The horizontal dashed line denotes the zero-vaccine efficacy. The efficacy estimates against hospitalization among seropositive due to DENV-4, among seronegative due to DENV-2 and DENV-4, were not estimable due to zero incidence among individuals in vaccine group. All the data have been reported in *(12)*.

### 2. Estimation of annual force of infection

The fraction of people who remain susceptible (0-infection) in age group $a$, during year $X$, is given by:

$$s_{a,X} = exp\left(-\sum_{i=0}^{a}\sum_{k=1}^{4}\lambda_{X-i}^{k}\right)$$

We assume that the force of infection (FOI) is time not serotype-specific, i.e. $\lambda_{X-i}^{k} = \lambda_{X-i}(k = 1,2,3,4)$, then the equation becomes

$$s_{a,X} = exp\left(-4\sum_{i=0}^{a}\lambda_{X-i}\right)$$

Now the fraction of people who experienced at least one infection is given by,

$$\pi_{a,X} = 1 - s_{a,X}$$

The fraction of people who has experienced only one infection is given by,

$$\phi_{a,X} = 4\left(1 - exp\left(-\sum_{i=0}^{a}\lambda_{X-i}\right)\right)exp\left(-3\sum_{i=0}^{a}\lambda_{X-i}\right)$$

Finally, the fraction of people who has experienced more than one infection is given by,

$$\alpha_{a,X} = 1 - s_{a,X} - \phi_{a,X}$$

We use the sero-prevalence data obtained from sero-surveys done in 2013 in Singapore. We have the number of participants and number of sero-positives for the age group 16–71.

The age group distribution in the sero-survey was: 16–20, 21–25, 26–30, 31–35, 36–40, 41–45, 46–50, 51–55, 56–60, and 60+ years. The data is obtained from (13).

A binomial log-likelihood is assumed for the FOI. The *optim* function in R used to find the maximum likelihood estimate of the FOI using the following equation:

$$\mathcal{L}(\lambda_{X-i}) = \sum_a \left(N_{a,X} - P_{a,X}\right)\left(log\left(1 - \pi_{a,X}\right)\right) + P_{a,X} log\left(\pi_{a,X}\right),$$

where, $N_{a,X}$ is the total number of individuals in age group $a$, and $P_{a,X}$ is the number of seropositive individuals.

The average annual force-of-infection in respective periods 2008–2013, 2003–2007, ..., 1953–1957 are denoted by $\lambda^p$, $p = 1,2, \dots ,12$. $\lambda^{13}$ represents the average annual FOI estimated for the years 1922–1952. The model fit with age-specific sero-prevalence data is presented in Fig S2.

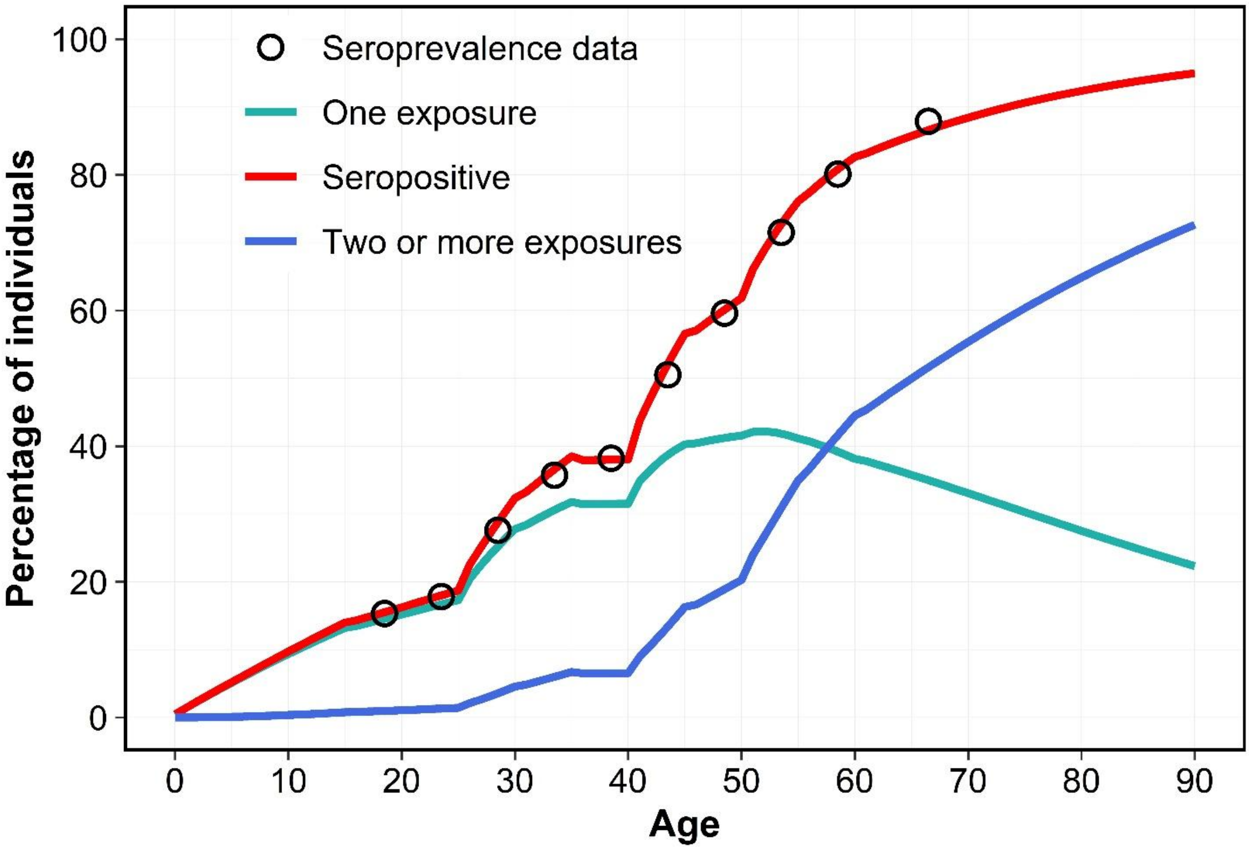

**Fig S2:** The percentage of population with different exposure history with age. The solid lines are calculated from catalytic model and the black circles are age-specific sero-prevalence data, form the survey conducted in 2013 in Singapore.

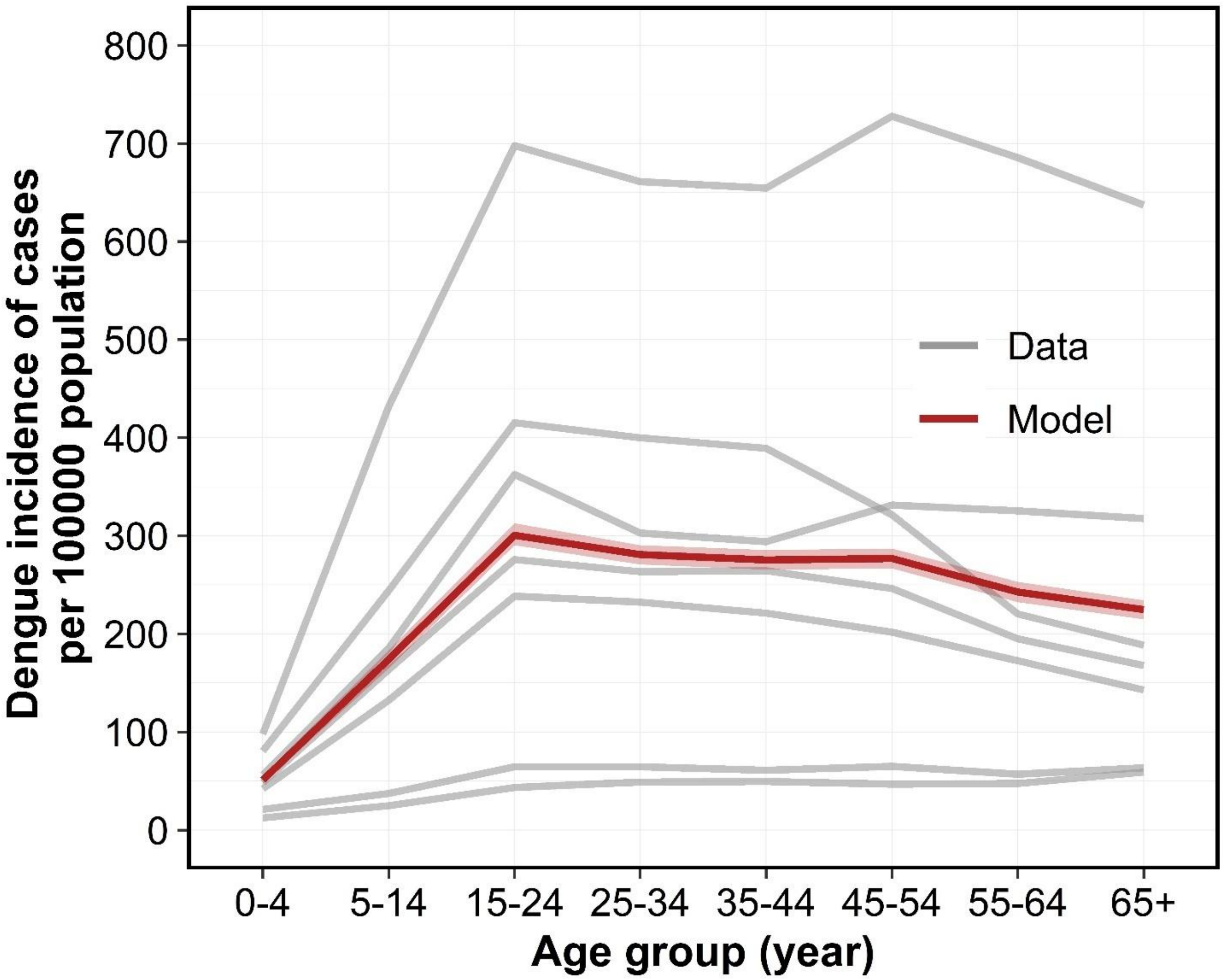

**Fig S3:** Model fitting with data. The gray lines are the incidence of cases per 100,000 population for each age group reported during 2014–2020. The dark red line is the median of incidence of cases per 100,000 population generated by the transmission model and the shaded region is the 95% credible interval.

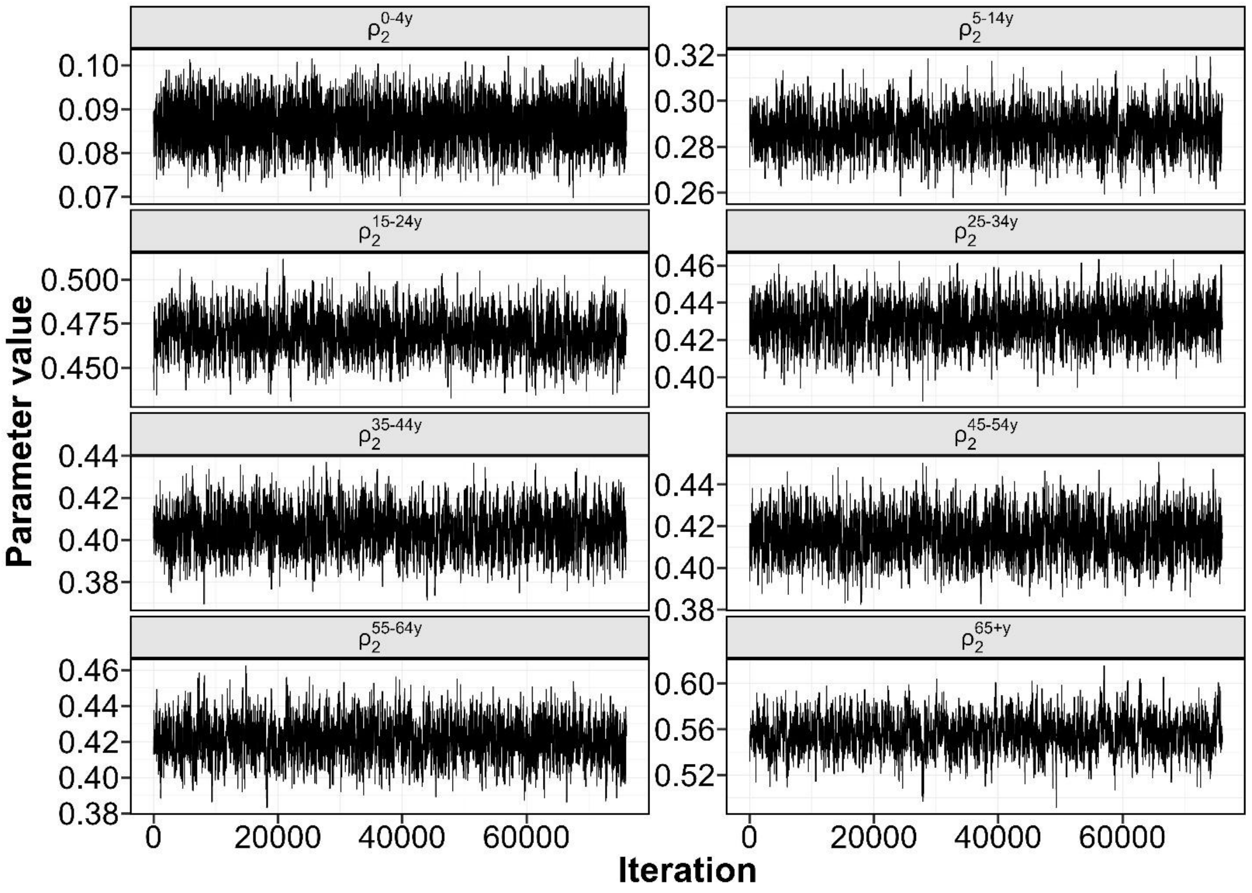

**Fig S4:** Trace plot of the MCMC chain of the estimated parameters.

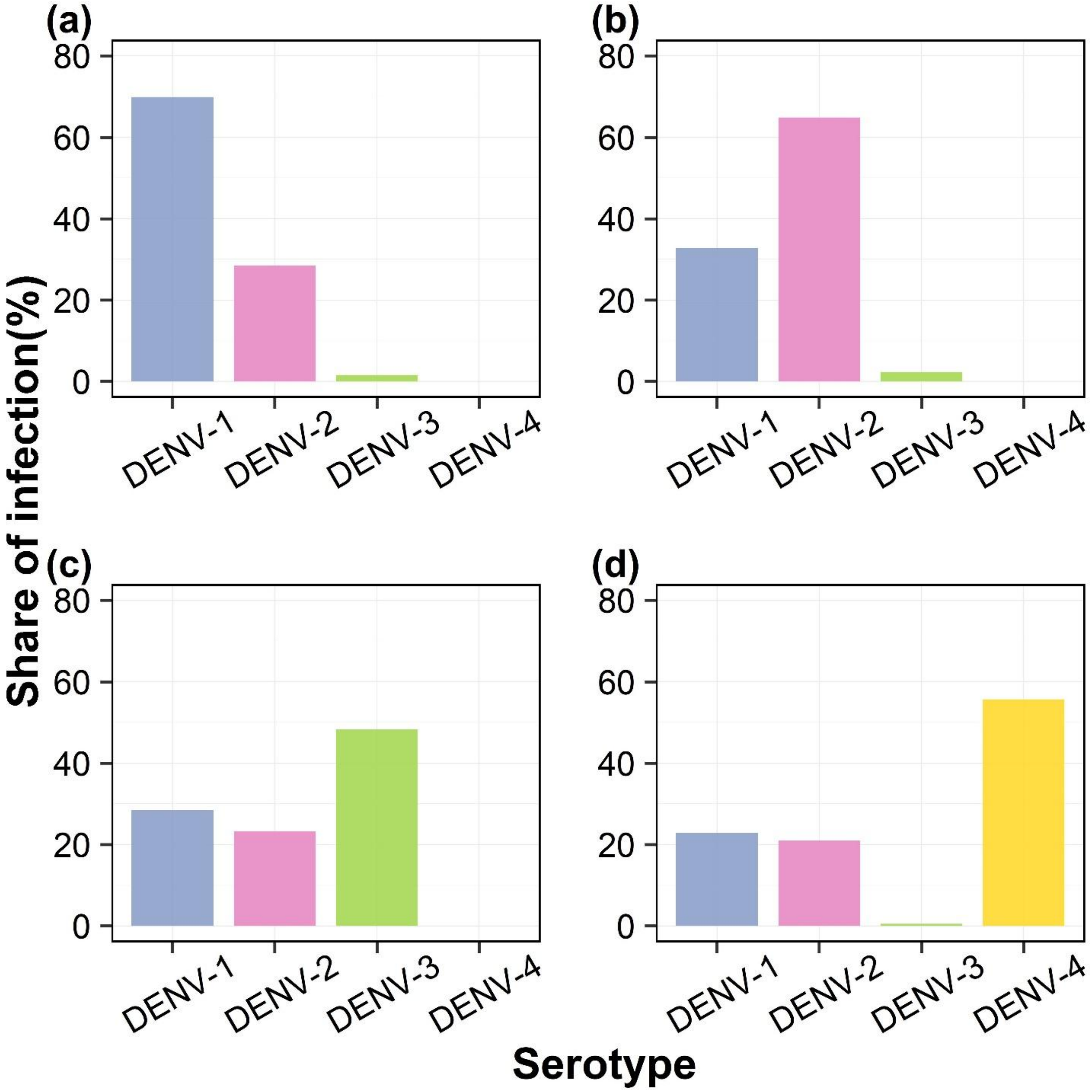

**Fig S5:** Serotype distribution of dengue infection during the rollout of vaccine for four serotype-dominant scenarios: (a) DENV-1, (b)DENV-2, (c) DENV-3, and (d) DENV-4, for baseline scenario (without vaccination).

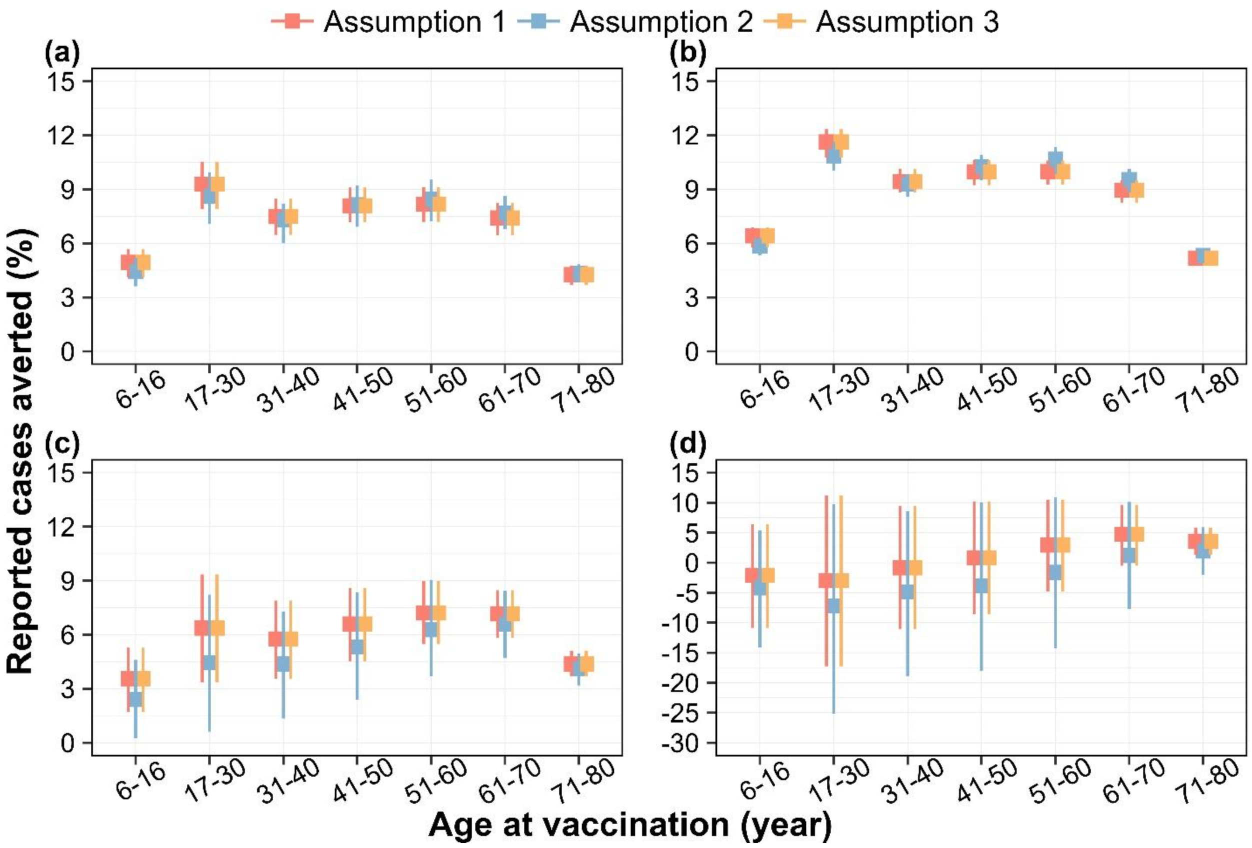

**Fig S6:** Percentage of dengue cases averted by targeted age groups, over a 10-year routine vaccination program with Qdenga under (a) DENV-1, (b) DENV-2, (c) DENV-3, and (d) DENV-4 dominant scenarios, for three different assumptions. Assumption 1 is the baseline assumption (presented in the main text) that both age-specific reporting rate and hospitalization rates depend on the pre-exposure history of infection, reporting rate among secondary infection is twice of that of primary infection; the hospitalization rate among secondary infection is 4 times higher than that of primary infection. In assumption 2, both reporting rate and hospitalization rate are only age-stratified but does not depend on pre-exposure history. In assumption 3, the reporting rate follows the same assumption as in assumption 1 but the hospitalization rate does not depend on pre-exposure history. All the estimates presented are for vaccine coverage 80%. Solid squares represent the mean model projections, and error bars indicate the corresponding 95% range of simulations.

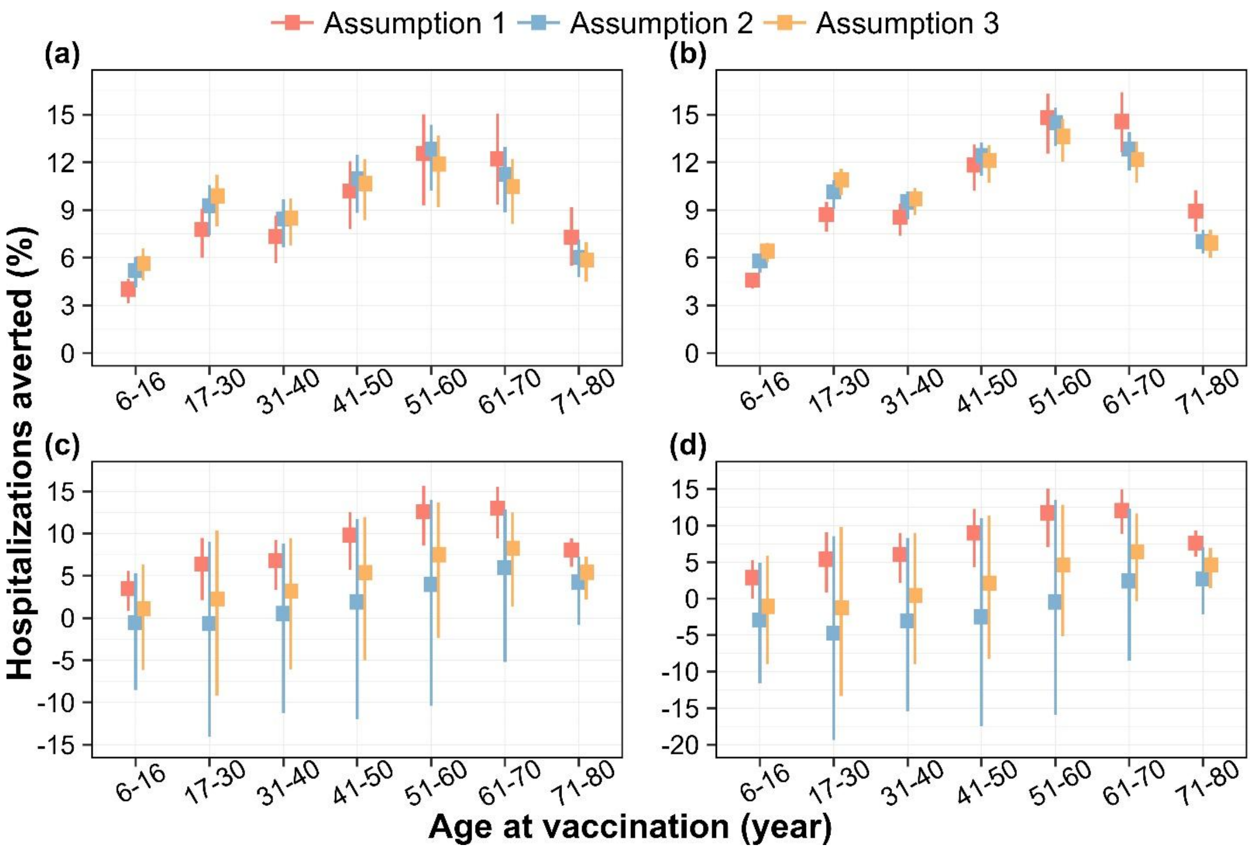

**Fig S7:** Percentage of hospitalizations averted by targeted age groups, over a 10-year routine vaccination program with Qdenga under (a) DENV-1, (b) DENV-2, (c) DENV-3, and (d) DENV-4 dominant scenarios, for three different assumptions. Assumption 1 is the baseline assumption (presented in the main text) that both age-specific reporting rate and hospitalization rates depend on the pre-exposure history of infection, reporting rate among secondary infection is twice of that of primary infection; the hospitalization rate among secondary infection is 4 times higher than that of primary infection. In assumption 2, both reporting rate and hospitalization rate are only age-stratified but does not depend on pre-exposure history. In assumption 3, the reporting rate follows the same assumption as in assumption 1 but the hospitalization rate does not depend on pre-exposure history. All the estimates presented are for vaccine coverage 80%. Solid squares represent the mean model projections, and error bars indicate the corresponding 95% range of simulations.

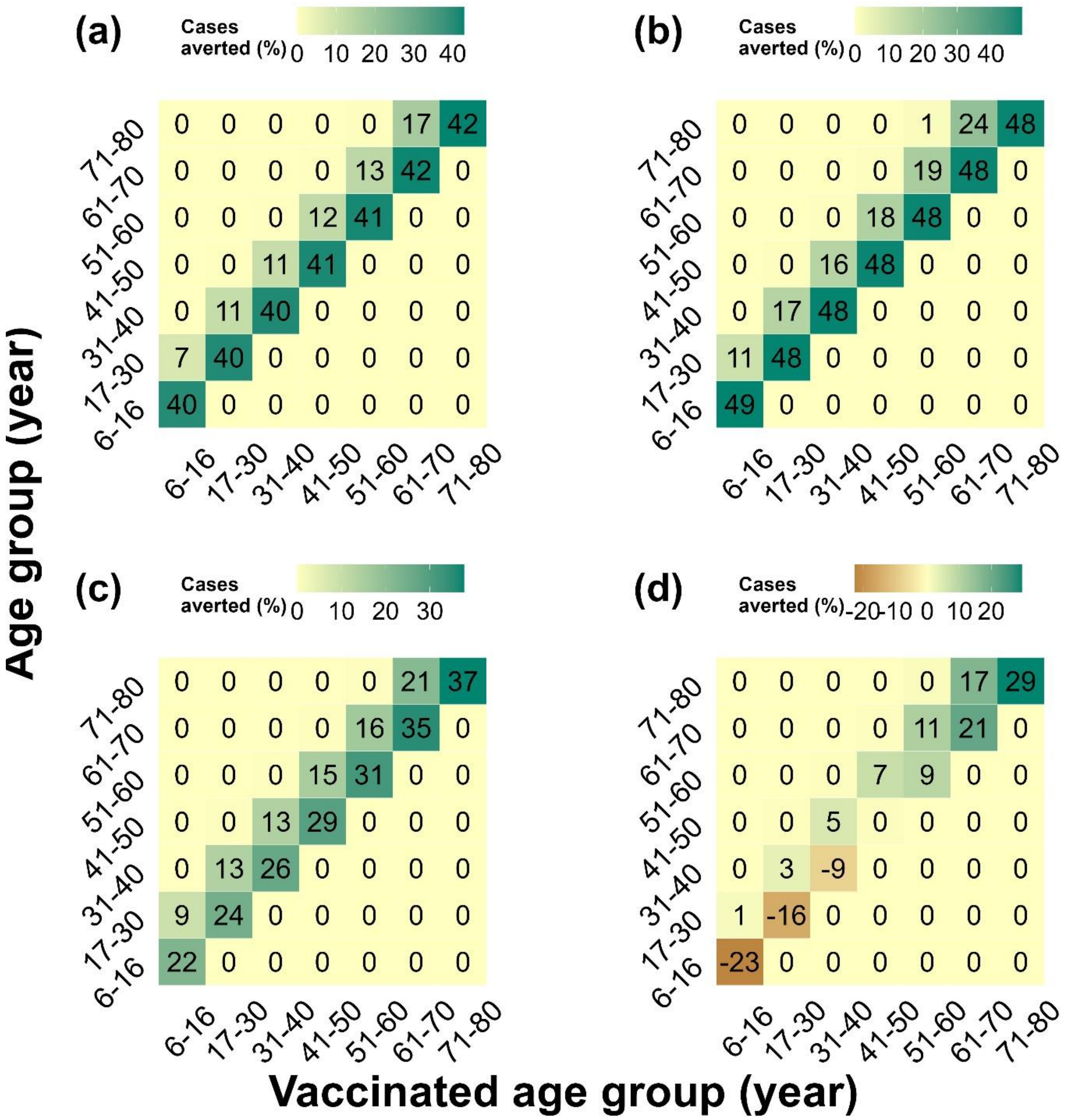

**Fig S8:** Mean estimate of impact of vaccination on different age-groups in terms of percentage averted reported cases in each age groups, under (a) DENV-1, (b) DENV-2, (c) DENV-3, and (d) DENV-4 dominant scenarios. The horizontal axis denotes different targeted age groups, and the vertical axis denotes the age groups for which the impact has been estimated. All the estimates presented are for vaccine coverage 80%.

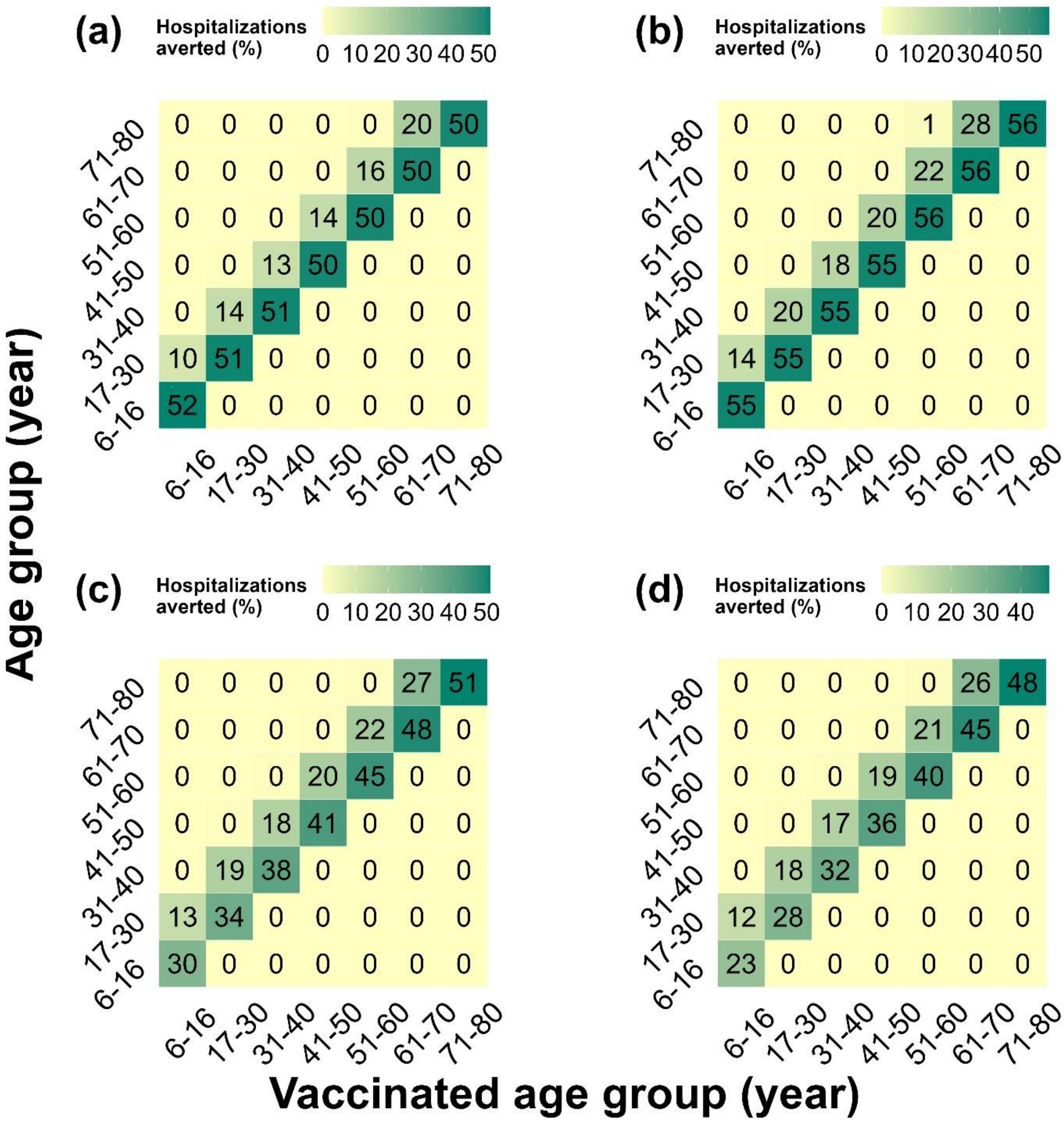

**Fig S9:** Mean estimate of impact of vaccination on different age-groups in terms of percentage of averted hospitalizations in each age groups, under (a) DENV-1, (b) DENV-2, (c) DENV-3, and (d) DENV-4 dominant scenarios. The horizontal axis denotes different targeted age groups, and the vertical axis denotes the age groups for which the impact has been estimated. All the estimates presented are for vaccine coverage 80%.

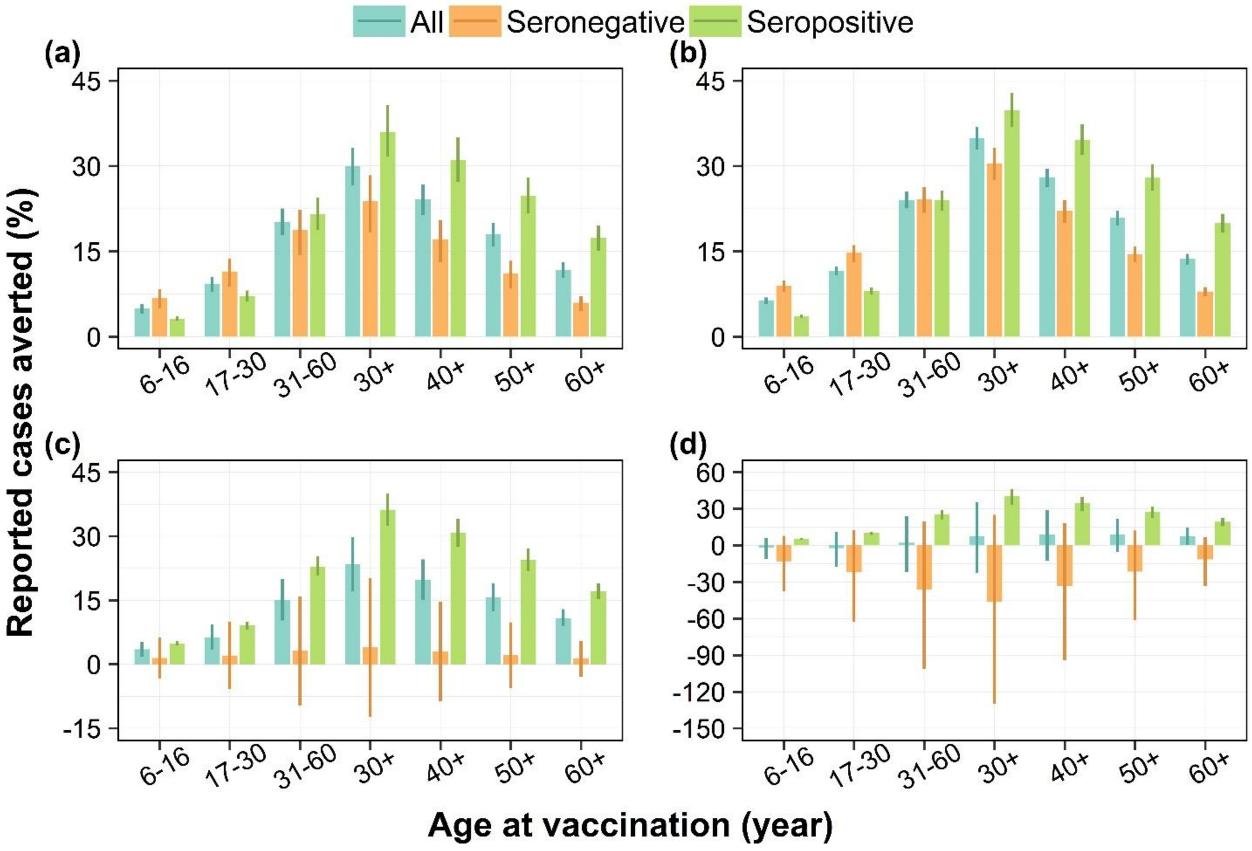

**Fig S10:** Percentage of reported dengue cases averted by targeted age groups over a 10-year routine vaccination program under (a) DENV-1, (b) DENV-2, (c) DENV-3, and (d) DENV-4 dominant scenarios, for the whole population, seropositive individuals only, and seronegative individuals only, assuming 80% vaccine coverage. Bars represent mean model estimates, and error bars indicate the corresponding 95% range of simulations.

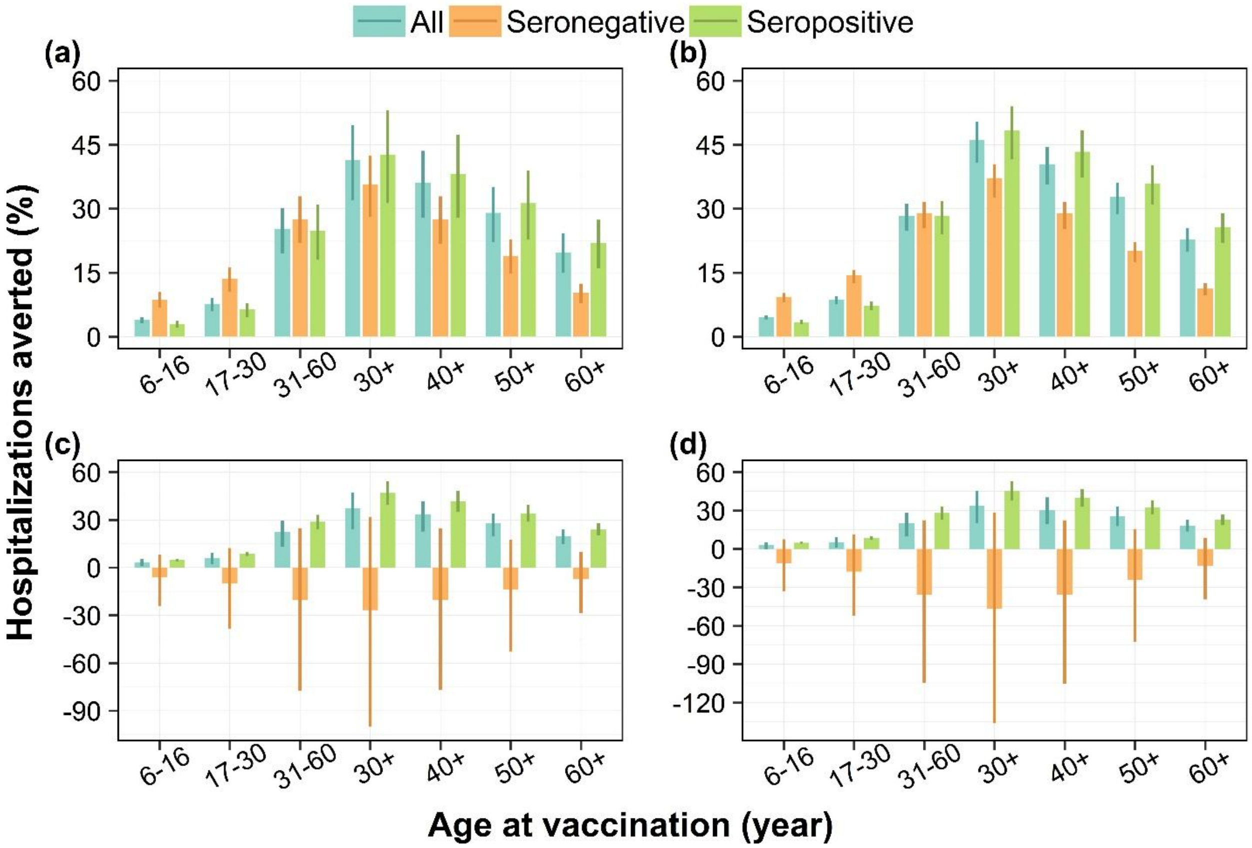

**Fig S11:** Percentage of reported hospitalizations due to dengue averted by targeted age groups over a 10-year routine vaccination program under (a) DENV-1, (b) DENV-2, (c) DENV-3, and (d) DENV-4 dominant scenarios, for the whole population, seropositive individuals only, and seronegative individuals only, assuming 80% vaccine coverage. Bars represent mean model estimates, and error bars indicate the corresponding 95% range of simulations.

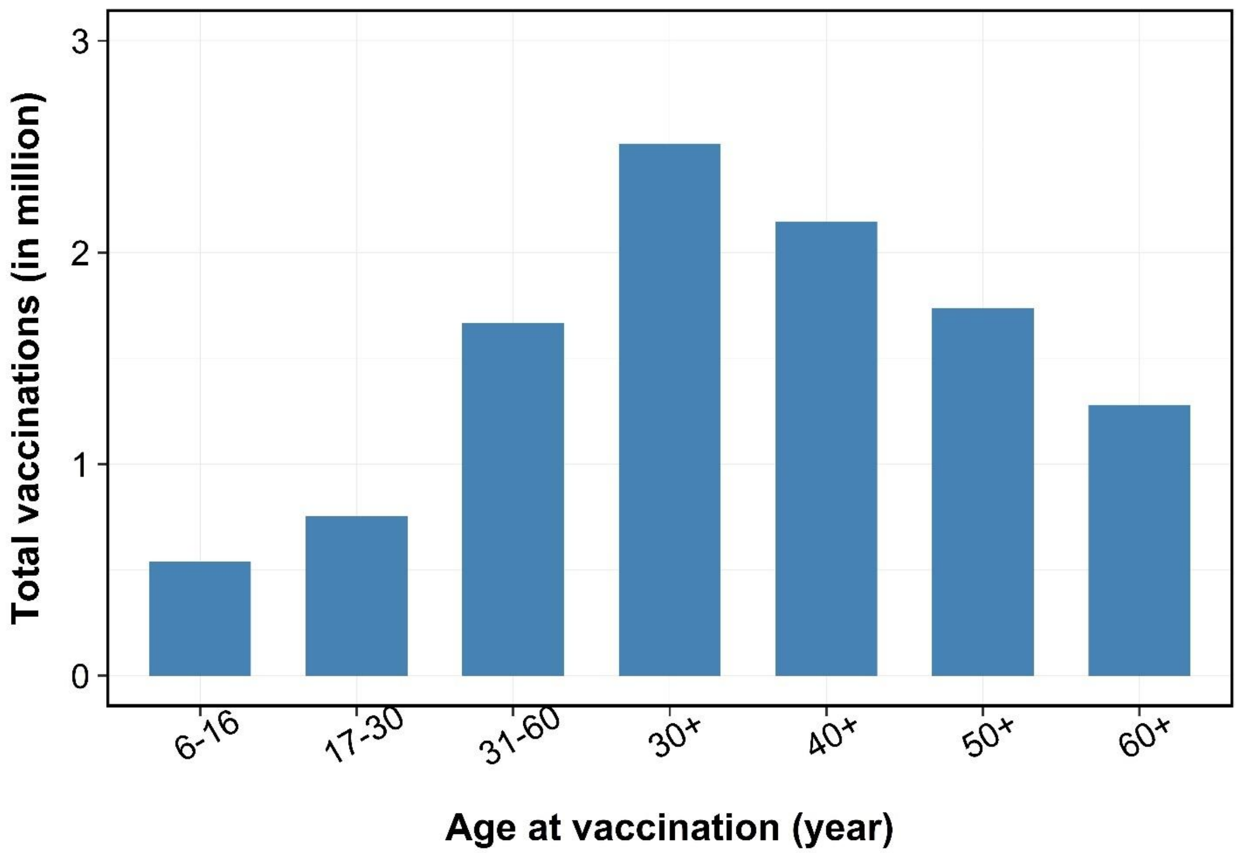

**Fig S12:** Total number of vaccinations for different targeted age groups. In each of the targeted age groups the coverage is 80%.
